# Physical therapy in patients with Parkinson’s disease treated with Deep Brain Stimulation: a Delphi panel study

**DOI:** 10.1101/2024.09.20.24314037

**Authors:** M. Guidetti, S. Marceglia, T. Bocci, R. Duncan, A. Fasano, K.D. Foote, C. Hamani, J.K. Krauss, A. A. Kühn, F. Lena, P. Limousin, A.M. Lozano, N.V. Maiorana, N. Modugno, E. Moro, M.S. Okun, S. Oliveri, M. Santilli, A. Schnitzler, Y. Temel, L. Timmermann, V. Visser-Vandewalle, J. Volkmann, A. Priori

## Abstract

Although deep brain stimulation of the subthalamic nucleus (STN-DBS) induces motor benefits in people with Parkinson’s disease (PwPD), the size and duration of the effects of STN-DBS on motor axial (e.g., postural instability, trunk posture alterations) and gait impairments (e.g., freezing of gait – FOG) are still ambiguous. Physical therapy (PT) effectively complements pharmacological treatment to improve postural stability, gait performance, and other dopamine-resistant symptoms (e.g. festination, hesitation, axial motor dysfunctions, and FOG) in PwPD who are non-surgically treated. Despite the potential for positive adjuvant effects of PT following STN-DBS surgery, there is a paucity of science available on the topic. In such a scenario, gathering the opinion and expertise of leading investigators worldwide was pursued to study motor rehabilitation in PwPD following STN-DBS. After summarizing the few available findings through a systematic review, we identified clinical and academically experienced DBS clinicians (n=21) to discuss the challenges related to PT following STN-DBS. A 5-point Likert scale questionnaire was used and based on the results of the systematic review along with a Delphi method. Thirty-nine questions were submitted to the panel – half related to general considerations on PT following STN-DBS, half related to PT treatments. Despite the low-to-moderate quality, the few available rehabilitative studies suggested that PT could improve dynamic and static balance, gait performance and posture. Similarly, panellists strongly agreed that PT might help in improving motor symptoms and quality of life, and it may be possibly prescribed to maximize the effects of the stimulation. The experts agreed that physical therapists could be part of the multidisciplinary team taking care of the patients. Also, they agreed on prescribing of conventional PT, but not massage or manual therapy. Our results will inform the rehabilitation and the DBS community to engage, publish and deepen this area of research. Such efforts may spark guidelines for PT following STN-DBS.

**GRAPHICAL ABSTRACT:** 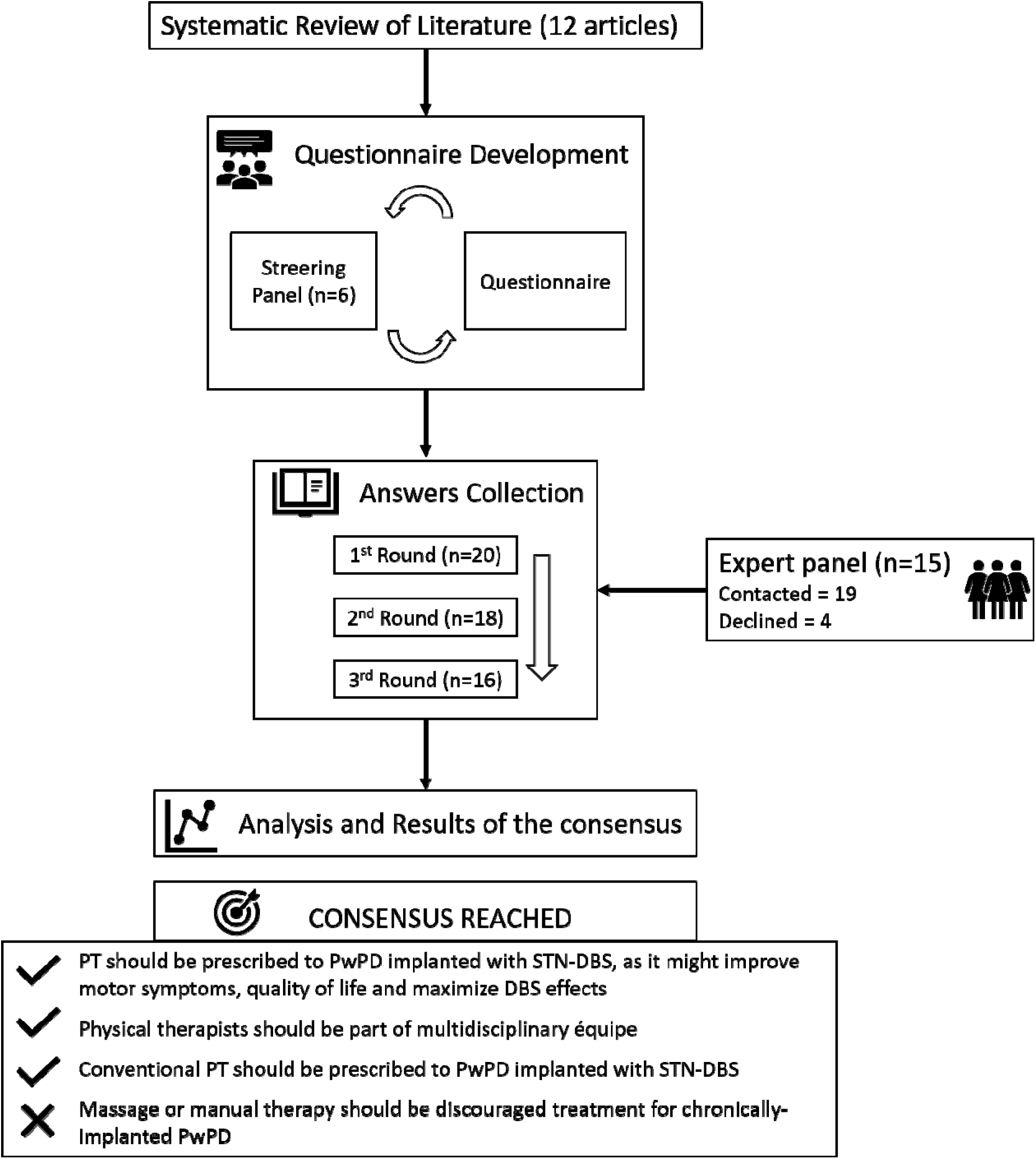

## 1. INTRODUCTION

Deep brain stimulation of the subthalamic nucleus (STN-DBS) is an established treatment for Parkinson’s disease (PD) (1,2), with a number of clinical studies suggesting long-term improvement of symptoms like tremor, rigidity, and akinesia (1,3). Nevertheless, STN-DBS is a symptomatic treatment, and the effect of stimulation on motor axial (e.g., postural instability, trunk posture alterations) and gait impairments (e.g., freezing of gait – FOG) is still unclear (4). Patients might experience no effect or even a worsening of these symptoms (4). This may be in part because stimulation parameters are often optimal for appendicular symptoms (1). Interestingly, some authors claimed that DBS has created a new phenotype of PD, with improvements in tremor, rigidity, bradykinesia, on–off fluctuations and dyskinesias, but progressive deterioration of gait, postural stability, speech and cognition (5,6).

Physical therapy (PT) is currently included in the multidisciplinary treatment of PD, but not specifically for patients treated with DBS (7,8). PT aims to optimize independence, safety, well=:Jbeing, and ultimately quality of life (9,10) with systematic reviews and meta-analysis confirming the PT-related improvement of motor and non-motor PD impairments (11–15). In particular, PT effectively complements pharmacological treatment to improve postural stability (13,16,17), gait (18–20), and those symptoms resistant to dopaminergic replacement (e.g. festination, hesitation, axial motor dysfunctions, and FOG) (9,12,21,22) in patients with PD (PwPD). Also, rehabilitative motor training stimulates a number of neuroplasticity-related events in PwPD (16), including neuronal growth, synaptogenesis, neurotrophic factor expression, and neurogenesis (17,23–26). Therefore, PT has the potential to be an effective adjuvant treatment to optimize motor outcomes after STN-DBS surgery. However, PT after STN-DBS has not been systematically assessed so far. Although the current recommendations allow the return to exercise within weeks following surgery, there is no explicit indication for PT (18). In addition, rehabilitative care in clinical settings is led by personal expertise of physical therapists. Only some insights of safety and effectiveness are currently available, but the studies are characterized by poor methodological rigor and great variability in terms of PT treatment, population, and timepoints of assessments. As a consequence, no solid scientific knowledge (e.g., guidelines) is currently available on motor rehabilitation after DBS surgery - instead, patients with STN-DBS are frequently excluded from exercise trials (27–31).

In such scenario, the opinion of leading experts in DBS field would boost the opening of the field of motor rehabilitation in patients with PD and STN-DBS. To foster this effort, we first performed a systematic scoping review of the research articles assessing PT programs in PwPD treated with STN-DBS, and then identified internationally recognized clinical and academic DBS experts to discuss these aspects participating in a Delphi method-based study.

## 2. METHODS

In this work, we first performed a systematic scoping review to gather the current knowledge on PT protocols in PwPD with STN-DBS. Based on the collected results and on the European Physiotherapy Guideline for Parkinson’s Disease (32), we created a 5-point Likert scale questionnaire regarding the role of PT and PT interventions in PwPD with STN-DBS to be answered by clinical and academically experienced DBS clinicians.

### 2.1. Systematic Scoping Review

A systematic scoping review of clinical research articles was performed according to previously published guidelines (33,34), since this type of review allows for a broad overview of topics (35). Literature search was conducted in PubMed/MEDLINE, considering the following search keywords: (“deep brain stimulation” OR “DBS”) AND (“physiotherapy” OR “physical therapy” OR “motor rehabilitation” OR “rehabilitation” OR “training” OR “exercise”) AND (“Parkinson’s disease” OR “PD”). We considered only clinical studies on PwPD with STN-DBS written in English and published from January 1^st^, 1994, to June 30^th^, 2024. Reviews, protocols, simulations studies, conference abstracts or editorials were excluded. Given the paucity of studies on the topic, we decided not to restrict further the inclusion criteria to be as inclusive as possible. After removing the duplicates, two independent reviewers (MG and NVM) screened the results of the search based on the titles and abstracts, and then evaluated the full texts of the selected articles. Conflicts were resolved by consensus, if necessary. Once publications that met the inclusion criteria were selected, the following data were extracted: author, year of publication, study design, characteristics of the subjects, DBS protocol and duration, PT protocol, outcomes and main results. Although the need for quality assessment of selected studies in scoping review has been questioned (36), some authors suggest it to improve clarity (37,38). Therefore, we decided to include the quality assessment of the included studies using the Downs and Black checklist (39), adjusted as previously published (40) to remove questions pertaining to a randomised controlled trial (see Table 1 in Supplementary Materials). Modified checklist assessed components of reporting, external and internal validity, assigning each article a final score and evaluation (total score: 11–13, excellent; total score: 9–10, good; total score: 7–8, fair; total score: ≤6, poor).

**Table 1.** Studies investigating physical therapy programs in patients with Parkinson’s disease and deep brain stimulation.

| Authors, year | Study design | Patients | DBS protocol and duration | PT protocol | Outcomes | Main results |
| --- | --- | --- | --- | --- | --- | --- |
| Cohen et al., 2007 (60) | Retrospective study | 73 patients (23 F; age, mean [range]: 60.6 [43-80] yo; disease duration mean [range]: 13.6 [3-27] yy; UPDRS - Part III: N.R. | Bilateral STN (n=71) and GPi-DBS (n=2)<br>N.R. duration<br>N.R. parameters | Multi-disciplinary personalized rehabilitation treatment (physical, occupational, and speech therapy + nutritional and psychological support) | UPDRS; FIM<br>Assessments pre- and post- hospitalisation | Significant improvements in motor performances and disability |
| Nampiarampil et al., 2008 (70) | Case series | Case 1: Male patient (age: 70 yo; disease duration: 7 yy; FIM: 39)<br>Case 2: Male patient with previous pallidotomy (age: 65 yo; disease duration: 15 yy; FIM: 25) | Case 1: bilateral DBS<br>Case 2: bilateral STN-DBS<br>N.R. duration<br>N.R. parameters | Case 1: physical, occupational and speech therapy (once a day, for 6 weeks).<br>Case 2: physical, occupational, and speech therapy (once a day, for 4 weeks). | FIM<br>Assessments pre- and post- hospitalization | Case 1: recovery of walking function with walker, and independency in ADL with assistance<br>Case 2: gait, tremor, and dyskinesia improved |
| Tassorelli et al., 2009 (67) | Pilot, pre-post, clinical study | 34 patients (15 F; age, mean ± SD: 57.6 ± 9.4 yo; disease duration, mean ± SD: 11.3 ± 4.4 yy; UPDRS - Part III, mean ± SD: 26.8 ± 12.8) | bilateral STN-DBS<br>n=13: <1 month after surgery;<br>n=8: 1-12 months after surgery;<br>n=13, >12 months after surgery<br>N.R. parameters | Personalized protocol:<br>Cardiovascular warm-up (5–10 min);<br>Stretching - trunk and limbs (15 min);<br>Strengthening muscles in a functional context - active-assisted or active isometric and isotonic exercises for trunk and limbs (10–15 min);<br>Relaxing muscles - especially for the flexor muscles (10 min);<br>Motor skills, coordination, and dual task performance (10 min); | UPDRS - Part III; FIM; mBI; MGHFAC; standing balance index.<br>Assessments pre- and post-rehabilitative intervention | Significant improvement of motor performance, functional independence, standing balance and independent walking ability. |
|  |  |  |  | Balance (10 min);<br>Gait training - with sensory cues (30 min).<br>Once a day, 5 days a week for 4-to-8 consecutive weeks |  |  |
| Nardo et al., 2014 (63) | Pilot, pre-post, clinical study | 9 patients (2 F; age, mean ± SD: 66.44 ± 5.7 yo; disease duration, mean ± SD: 12.2 ± 6 yy; UPDRS - Part III, mean ± SD: 36.7 ± 6.4) | DBS (months after surgery, mean ± SD: 3.11 ± 1.19) N.R. parameters | Protocol comprising:<br>Body weight supported and robotic-assisted treadmill training: speed at 1.5 km/h, increased up to 3 km/h as tolerated (45 min)<br>Once a day, for 5 weeks | UPDRS - Part III; Gait kinematics, kinetic, and spatiotemporal parameters<br>Assessments pre- and post-rehabilitative intervention | Significant improvements in gait performance, in all the spatiotemporal gait parameters, and in maximal ankle plantar flexion angle in the toe-off phase |
| Luna et al., 2018 (65) | Cross-over clinical trial | 12 patients (5 F; age, mean ± SD: 61.5 ± 10.4 yo; disease duration, mean ± SD: 18.6 ± 5.2 yy; mH&Y, mean ± SD: 2.3 ± 0.3) | bilateral STN-DBS (months after surgery, mean ± SD: 1.7 ± 0.6) N.R. parameters | EG: treadmill training with body weight support (30min) + physical therapy (60min)<br>CG: treadmill training without body weight support (30min) + physical therapy (60min)<br>Treadmill training: speed at 0.5 km/h, increased by increments of 0.5 km/h as tolerated.<br>Physical therapy: stretching exercise for trunk, upper and lower limbs muscles (2min); strengthening exercises for upper, lower limbs, trunk and scapular muscles (for each, 3 sets of 15 repetitions); exercise for balance (bipodal, tandem and unipodal stance – 2 sets of each)<br>twice a week for 8 weeks | Gait kinematics, spatiotemporal and angular parameters<br>Assessments pre- and post-rehabilitative intervention | Significant improvements in pelvis' range of motion; hip's range of amplitude; knee flexion on swing phase; and foot progression range of motion (EG group) |
| Bestaven et al., 2019 (66) | Pilot, pre-post, clinical study | 10 patients (3 F; age, mean ± SD: $67.6 \pm 6.3$ yo; disease duration, mean ± SD: $18.8 \pm 4$ yy; UPDRS - Part III: N.R.) | bilateral STN-DBS (months after surgery, mean ± SD: $94.8 \pm 37.2$, 60–175 Hz; 60–90 μs; 2.1–4.6 V) | Protocol comprising:<br>Stretching exercise for trunk muscles (75 min);<br>Strengthening exercises for trunk muscles, in extension, flexion and rotation (75 min);<br>Cardiovascular training (30 min)<br>twice a day, 5 days a week for 4 weeks | UPDRS - Part III; UPDRS - Part III axial score (items 18, 19, 20, 22, 27–30); UPDRS - Part III gait score (item 30); UPDRS - Part III postural instability score (item 29); ABD; BBS; 3D kinematic gait analyses<br>Assessments pre- and post-rehabilitative intervention | Significant improvements in gait performances and posture; significant decrease in daily number of falls |
| Sato et al., 2019 (61) | Retrospective study | 16 patients (5 F; age, median ± IQR: $61.5 \pm 9.5$ yo; disease duration, median ± IQR: $13 \pm 8$ yy; UPDRS - Part III, median ± IQR: $17.5 \pm 7.75$) | STN-DBS (median: 130 Hz; 60 μs; 1.68 V) N.R. duration | Protocol aiming to improve muscle strength, flexibility, balance, and gait.<br>Flexibility: active assistive range of motion exercise for ankle, hip, and trunk joints<br>Strength and balance: dynamic balance exercise in the quadrupedal (cat and dog, diagonal balancing exercise) and standing positions (toe-heel weight bearing, one-leg standing, step position)<br>Gait: active assistive gait training<br>40min a day, for 14 days | Mini-BESTest; TUG; UPDRS-III; BI<br>Assessments before, three days after and 2 weeks after surgery | Significant improvements in balance and gait ability |
| Naro et al., 2020 (64) | Case-controlled pilot study | EG: 10 patients with STN-DBS (4 F; age, mean ± SD: $62 \pm 5$ | bilateral STN-DBS (months after surgery: | EG, CG: RAS-assisted treadmill training (30min) + physical therapy (60min) | UPDRS – part III; TUG; 10MWT; BBS; FES; ACE-R; EEG | EG: Significant improvements in motor performance |
|  |  | yo; disease duration, mean ± SD: $15 \pm 2$ yy; UPDRS - Part III, mean ± SD: $19.1 \pm 9.03$)<br>CG: 10 patients without DBS (5 F; age, mean ± SD: $62 \pm 4$ yo; disease duration, mean ± SD: $14 \pm 2$ yy; UPDRS - Part III, mean ± SD: $27.54 \pm 1.12$) | >12; 130–240 Hz; 60–120 μs; 2.2–3.6 V) | RAS-assisted treadmill training: bpm at $85 \pm 5$ (0.43 m/s), increased by 5 bpm every 3 min up to 120 bpm (0.61 m/s).<br>Physical therapy: exercises to improve flexibility, balance, gait, and muscular tone and resistance.<br>once a day, 6 days a week, for 4 weeks | Assessments pre- and post-rehabilitative intervention | (self-confidence in balance, sit-to-stand, velocity), walking (velocity), and remodulation of gait cycle-related beta oscillations. Both groups: significant improvements in dynamic and static balance, cognitive performance, and the fear of falling |
| Li et al., 2020 (68) | Pilot, clinical study | 16 patients (8 F; age, mean ± SD: $60.25 \pm 5.6$ yo; disease duration, mean ± SD: $10.38 \pm 4.33$ yy; MDS-UPDRS - Part III, mean ± SD: $59.38 \pm 17.07$) | bilateral STN-DBS<br>N.R. duration<br>N.R. parameters | Multi-disciplinary treatment (DBS, rehabilitation, medication, psychotherapy), comprising:<br>Core strength training;<br>Postural stability training;<br>Training of sensory function | PDQ-39; MDS-UPDRS - Part III; MDS-UPDRS 3.12; BBS; LoS.<br>Assessments pre- and post-surgery, 6 months post-surgery, 12 months post-surgery | Significant improvements in QoL, motor and balance performance at 6 and 12 months |
| Liang et al., 2020 (69) | Pilot, clinical study | 15 patients (8 F; age, mean ± SD: $62.5 \pm 8$ yo; disease duration, mean ± SD: $10.5 \pm 4.47$ yy; MDS-UPDRS - Part III, mean ± SD: $55.06 \pm 16.77$) | bilateral STN-DBS (130–170 Hz; 60–90 μs; 1.5–3.5 V)<br>N.R. duration | Protocol comprising:<br>Stretching exercises for neck, shoulders, chest, and waist muscles (10 min);<br>Strengthening of back, posterior shoulder, gluteal muscles (at least 1 set of 10 to 15 repetitions for each);<br>Back extension and bridge exercise (5 sec each);<br>Education to the patient<br>once per day for 8 weeks | PDQ-39, MDS-UPDRS III, degree of camptocormia<br>Assessments pre-, at 1 month and 6 months after surgery | Significant improvements in camptocormia |
| Sato et al., 2022 (62) | Pre-post, clinical study | 60 patients (28 F; age, mean ± SD: $60.7 \pm 8.9$ yo; disease duration, mean ± SD: $12.2 \pm 4.6$ yy; MDS-UPDRS - Part III, mean ± SD: $18.1 \pm 8.6$) | STN-DBS ($131.2 \pm 6.5$ Hz; $58.8 \pm 4.9$ μs; $1.8 \pm 0.5$ mA)<br>N.R. duration | General program combining muscle-strengthening exercises, stretching, and balance exercises.<br>40-60 min a day, for 14 days | Mini-BESTest; TUG; TIS; Lower Extremity Extension Torque; 10 Toe-Tapping Seconds; Postural Sway Test<br>Assessments before and three days after surgery, and just before discharge) | Significant improvements in physical function, balance, and gait ability |
| Canesi et al., 2024 (71) | Case-controlled pilot study | EG: 22 patients with DBS (9 F; age, median ± IQR: $63.5 \pm 13.5$ yo; disease duration, median ± IQR: $17 \pm 9$ yy; MDS-UPDRS, median ± IQR: $105.5 \pm 45.55$)<br>CG: 25 patients without DBS (9 F; age, median ± IQR: $69 \pm 11$ yo; disease duration, median ± IQR: $15 \pm 6$ yy; MDS-UPDRS, median ± IQR: $86 \pm 30$) | DBS (months after surgery, median ± IQR: $72 \pm 69.6$)<br>N.R. parameters | Multi-disciplinary treatment (occupational therapy, speech therapy), comprising physical therapy:<br>Morning session: warming-up (passive and active mobilization exercises for both upper and lower limbs - 10min), aerobic exercises (walking and cycling, with intensity between 50-80% of the maximal heart rate – 15min), active mobilization exercises and strengthening exercises (60–75% of the estimated 1RM - 15min), postural/proprioceptive exercises (10min), and cooling down (passive and active mobilization exercises – 10min)<br>Afternoon session: warming-up (10min), treadmill (15min), aerobic exercise (intensity between 50-80% of the maximal heart rate – 15min),<br>proprioceptive exercises<br>(15min) and cooling down<br>(passive and active<br>mobilization exercises –<br>10min)<br>60min, twice a day, 5 days a week<br>for 4 consecutive weeks | MDS-UPDRS; BBS; SPDDS; TUG; 6MWT; MoCA<br>Assessments pre- and 24h after rehabilitative intervention | EG and CG improved physical functioning and performance, balance function and independence in ADL, but without difference between groups |

F = females; yo = years old; yy = years; UPDRS = Unified Parkinson's Disease Rating Scale; STN-DBS = subthalamic nucleus deep brain stimulation; GPi-DBS = globus pallidus internus deep brain stimulation; DBS = deep brain stimulation; N.R. = not reported; FIM = Functional Independence Measure; BI = modified Barthel Index; MGHFAC = Massachusetts General Hospital Functional Ambulation Classification; EG = experimental group; CG = control group; ABD = Activities-specific Balance; BBS = berg balance scale; Mini-BESTest = Mini-Balance Evaluation Systems Test; TUG = Timed Up and Go; BI = Barthel Index; RAS = rhythmic auditory stimulation; 10MWT = 10 meters walking test; FES = falls efficacy scale; ACE-R = Addenbrooke's Cognitive Examination-Revised; PDQ-39 = Parkinson's Disease Questionnaire; MDS-UPDRS = Movement Disorder Society-Unified Parkinson's Disease Rating Scale; LoS = Limits of Stability; TIS = Trunk Impairment Scale; SPDDS = Self-Assessment Parkinson Disease Scale; 6MWT = 6 Min Walk Test; MoCA = Montreal Cognitive Assessment.

### 2.2. Questionnaire development

As proposed by Kerlinger et al., 1973 (41), the questionnaire was based upon an extensive review of the literature and the European Physiotherapy Guideline for Parkinson’s Disease (32). From the scoping review, we defined a taxonomy of the outcome measures, and related each of them to an improvement area, and a taxonomy of the PT proposed in published studies. Then, the concepts identified in the two taxonomies were translated into the two sections of the questionnaire: one, more general, focusing on the opportunity and potential benefits of PT for DBS patients; the other, focusing on the different PT treatments. In addition, guidelines (32) were used to include other PT treatments not covered by the literature review (see Table 2 in Supplementary Materials).

**Table 2.**
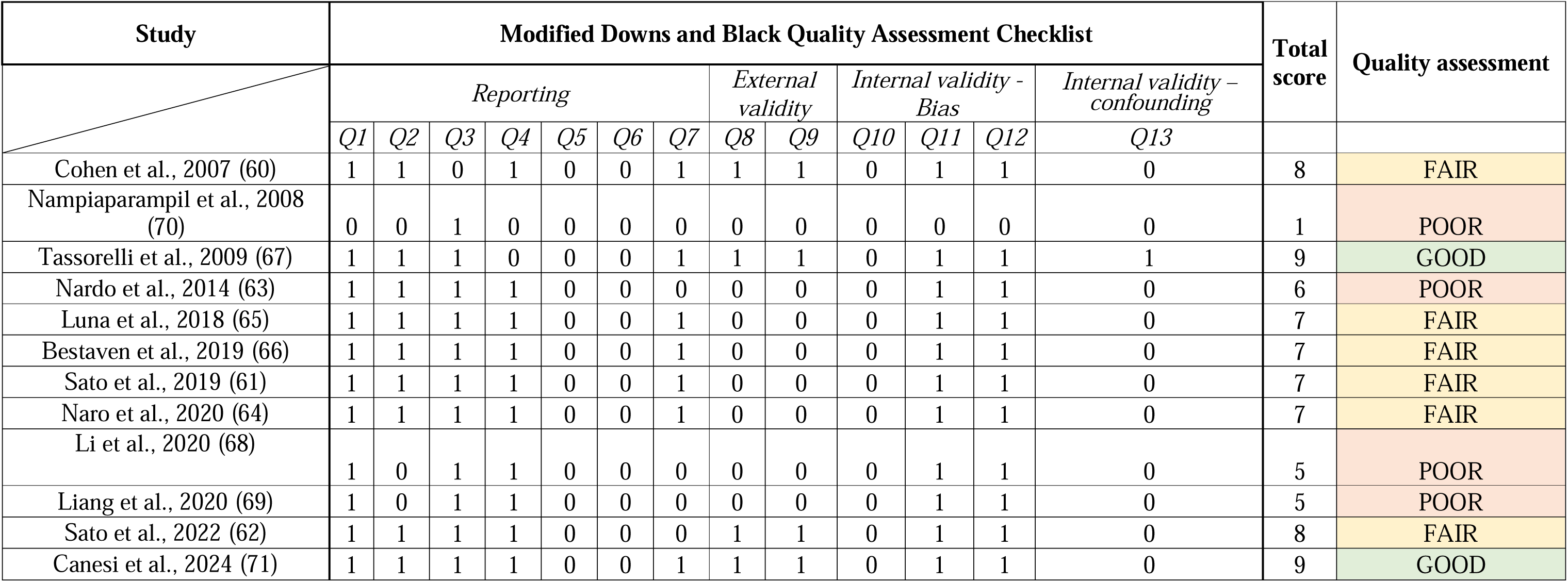
Modified Downs and Black Quality Assessment Checklist. (40)

### 2.3. Delphi Methodology

The Delphi technique is a multi-phase procedure that combines personal viewpoints into a general consensus within a group (panel) (42,43). In fact, the members of the group (panellists) anonymously complete a questionnaire multiple times (rounds), receiving aggregated results from each round each time (44–46), so that they can re-evaluate their judgments. This ensures the advantages of interacting groups (e.g., inclusion of people from diverse backgrounds) eliminating the disadvantages (e.g., the presence of dominant members) (47). For the purpose of our study, a modified Delphi process (41) was created in three rounds, which are thought to be adequate to gather the necessary data and come to a consensus (44,47–51). Following a thorough review of the literature, a Steering Committee (SC) of experts (n = 6) based on the collaborative network of the leading authors discussed the topics and, in accordance with the methodology outlined in Section 2.2, created a structured questionnaire using a five-point Likert scale (1=strongly disagree; 2=disagree; 3=undecided; 4=agree; 5=strongly agree) (52). In rounds one, two and three, the SC together with a broader Experts Panel (EP = 15) conducted quantitative assessments to reach the consensus. Since there is no precise standard for defining an “expert” (53), we chose to involve positional leaders in the scientific area, as previously proposed (54). We considered a response rate of >70% for each round to preserve the rigor of the technique (55). Electronic questionnaires were utilised in all steps of the process. To prevent confirmation bias, if a statement came to a consensus in either the first or second round, it was not included in the next round. Despite the absence of guidelines (53), we considered a “consensus reached” when >80% of the responses fell in the same response label (52,56). Descriptive statistics (median ± IQR) were used to analyze and report the data, as recommended (47,57–59). Also, to highlight the strength of support for each round, we reported the results of each round separately in both textual (i.e., with median ± IQR) and graphical representation (53).

## 3. RESULTS

### 3.1. Systematic scoping review

Our search yielded 632 articles (Fig. 1). Of those, 615 were excluded after reviewing titles and abstracts against inclusion/exclusion criteria, while 17 were further assessed as full paper for eligibility. Of these, only 12 met our inclusion criteria and therefore selected for our scoping review (60–69). The characteristics of the included studies are summarised in Table 1. One was a case series (70), seven were pilot clinical studies (62,63,65–69), two retrospective studies (60,61) and two case-controlled studies (64,71), for a total of 279 patients with DBS implant enrolled. Of these, 245 had STN-DBS (169 bilateral, 76 not specified), 2 had bilateral globus pallidus internus (GPi)- DBS, and 32 had DBS with no specified anatomical target. The number of participants per study ranged between 1 (70) and 73 (60) participants; eight studies involved 20 participants or less (61,63–66,68–70), while four more than 20 (60,62,67,71). The mean age of participants ranged from 57.6 (67) to 67.6 (66) years, with a mean baseline disease severity from 19.1 (UPDRS, part III) (64) to 105.5 (MDS-UPDRS, part III) (71) and a mean disease duration from 10.5 (69) to 18.8 (66). Only five studies reported the characteristics of the stimulation (61,62,64,66,69), and seven studies did not specified the duration of DBS treatment before PT treatment (60–62,64,68–70) - see Table 1. As for quality assessment, two studies (67,71) were classified as presenting good methodological quality, six (60–62,64–66) as fair, and four (63,68–70) as poor, according to the Modified Downs and Black Quality Assessment Checklist (Table 2). In general, the studies attended the criteria regarding the reporting section, however, a few studies (62,63,68–70) did not report the actual probability values of results, and none provided estimates of random variability for the main outcomes nor reported the characteristics of patients lost to follow-up. Due to limited sample size, external validity could not be guaranteed for most of the articles. As for internal validity, none clearly stated the potential use of data dredging nor considered drops-out of patients at follow-ups (except for (67)).

**Fig.1.**
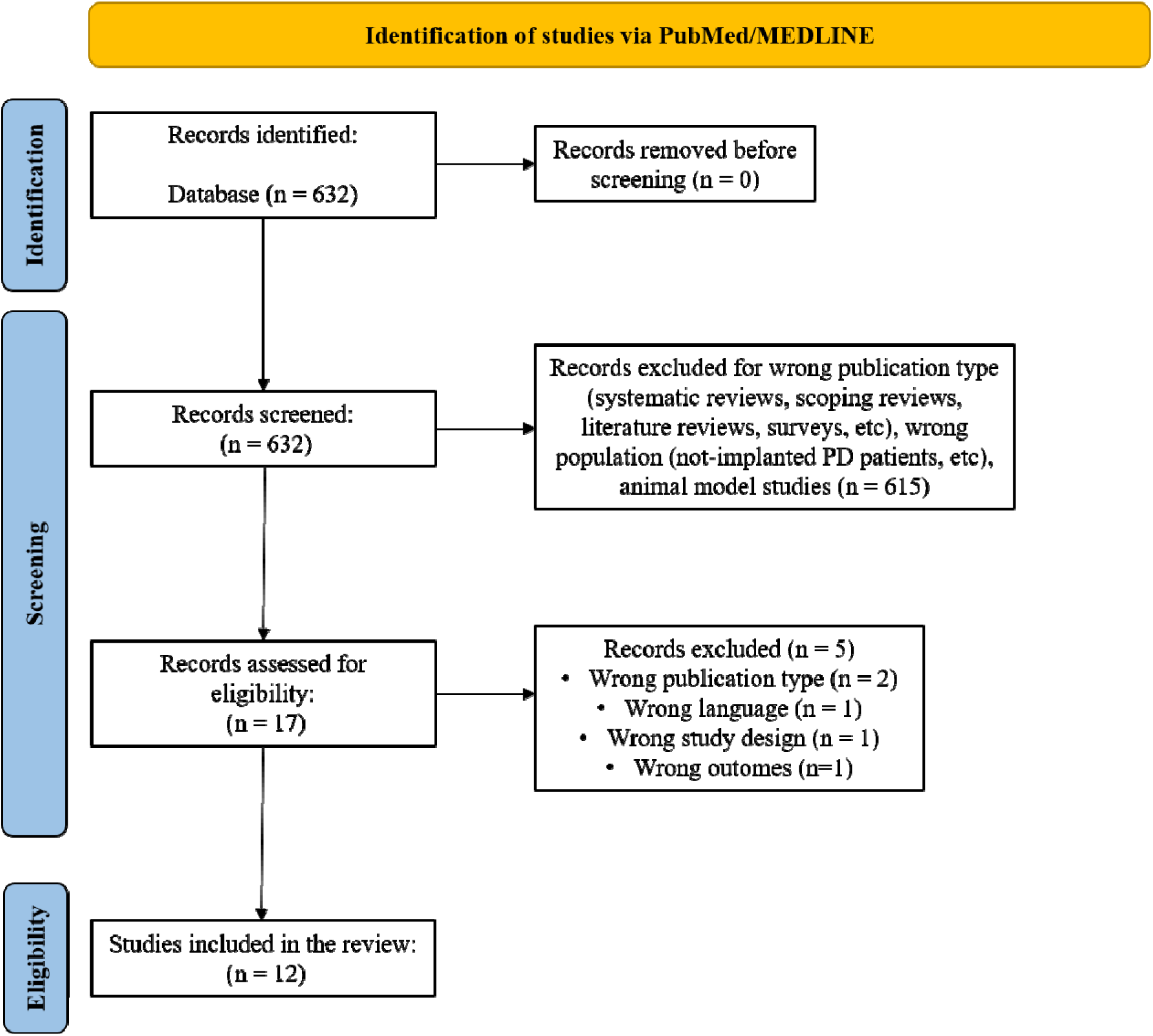
Flowchart of the scoping review selection process.

### 3.2. PT outcomes and areas of assessments

The effect of the PT interventions was evaluated through various outcomes across the studies (Table 1), which assessed both motor/functional, biomechanical (e.g., gait analyses) and neurophysiological (e.g., EEG) changes. The selected studies examined the role of PT in PwPD undergoing STN-DBS in 6 main areas of assessment: (I) Motor symptoms and motor decline, as assessed mainly through the UPDRS – part III, including its different scores (e.g., axial score and gait score), or Movement Disorder Society Sponsored Revision of the UPDRS (MDS-UPDRS)(60,61,63,64,66–69,71); (II) Gait performance, as assessed mainly though TUG (61,62,64,71) and gait analyses (63,65,66); (III) Balance and postural instability, as assessed mainly though BBS (64,66,68,71) and Mini-BESTest (61,62); (IV) Quality of life or activities of daily living, as assessed mainly though FIM (60,67,70) and PDQ-39 (68,69); (V) Timing of PT treatment, in terms of moths after the neurosurgery. Although half of the selected studies did not report the time between surgery and rehabilitation (60–62,68–70), three considered patients in chronic stimulation (e.g., several years) (64,66,71), while two patients with only few months of DBS (<one year) (63,65). One study (67) enrolled patients with different timings (67). As shown in Table 2 in Supplementary Materials, these areas of assessment were used to build the questionnaire for the Delphi panel.

### 3.3. PT treatments

PT treatments and protocols varied considerably across the selected studies (Table 1). Most of them studied the effect of aerobic training with mobility, stretching, strengthening, balance and gait exercises or a combination thereof (61,62,66,67,69), while four (60,68,70,71) considered a multidisciplinary approach, where physical therapy was a part of a more articulated rehabilitation care and associated to other interventions such as occupational or speech therapy. Of them, only one study (71) reported a clear description of the characteristics of the intervention. Three studies described the use of treadmill training; however, Nardo et al., 2017 (63) associated it with body weight and robotic support, Luna et al., 2018 (65) with body weight support and physical therapy (stretching, strengthening and balance exercises), and Naro et al., 2020 (64) with rhythmic auditory stimulation. Similarly, PT protocols remarkably differed in terms of intensity, frequency, and duration. Only three studies reported the intensity (i.e., session length) of the treatment (61,62,71), which ranged from 40 to 60min. The frequency ranged from twice weekly for eight weeks (65) to twice a day weekly for four weeks (66,71), for a total duration ranging from 2 (61,62) to 8 (65,67,69) weeks.

### 3.4. Delphi panel results

For the SC, 7 authors were invited but only 6 agreed to participate (SC=6, response rate: 85.7%). For the EP, out of the 20 authors identified, 2 declined to participate and 3 did not reply (EP=15, response rate: 75%). Therefore, the overall number of panellists was 21 (overall response rate: 77.7% - see Table 3), which is a number of experts within the recommended range (47,72).

**Table 3.**
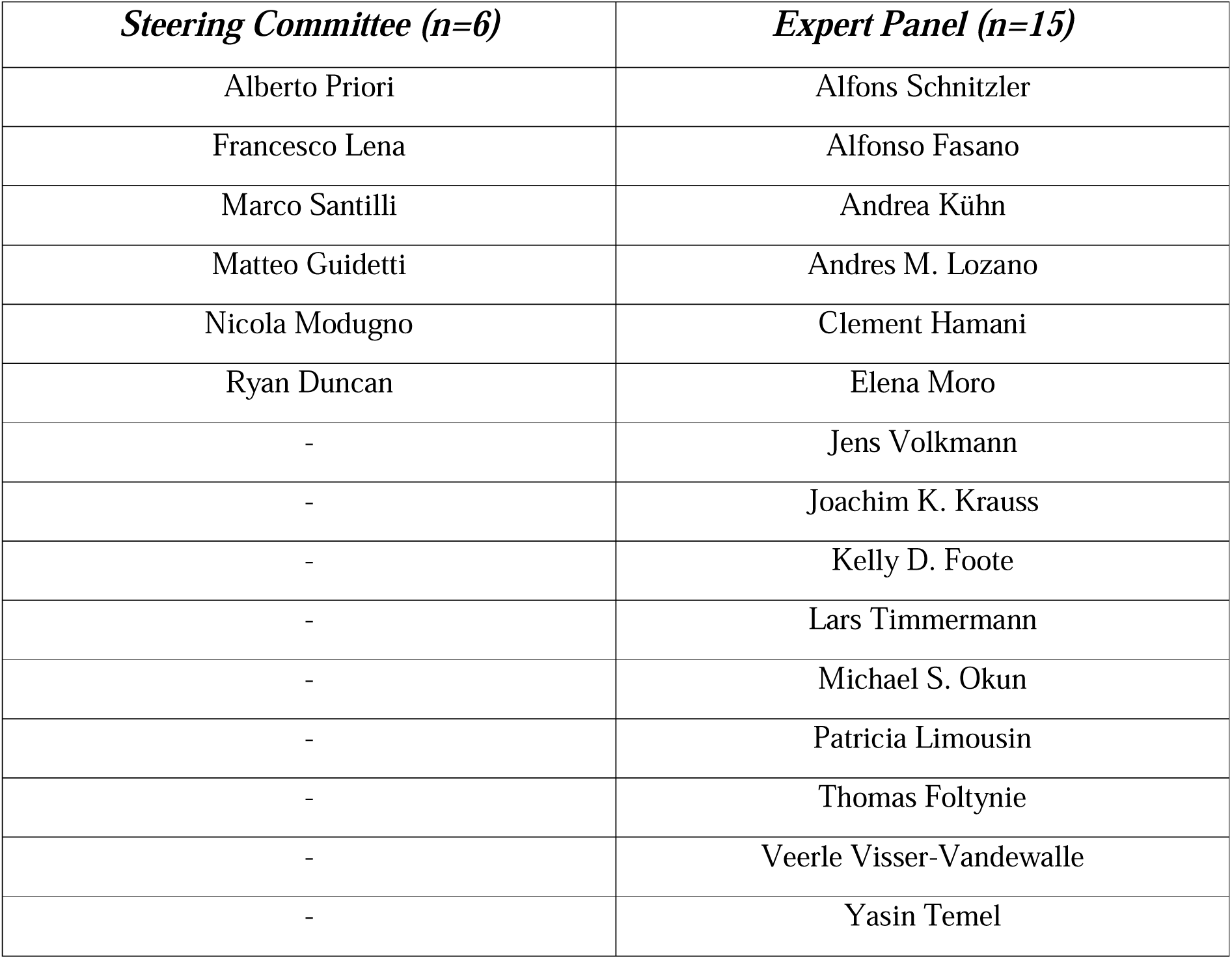
Authors panelists in the Steering Committee (SC) and the Expert Panel (EP).

| *Steering Committee (n=6)* | *Expert Panel (n=15)* |
| --- | --- |
| Alberto Priori | Alfons Schnitzler |
| Francesco Lena | Alfonso Fasano |
| Marco Santilli | Andrea Kühn |
| Matteo Guidetti | Andres M. Lozano |
| Nicola Modugno | Clement Hamani |
| Ryan Duncan | Elena Moro |
| - | Jens Volkmann |
| - | Joachim K. Krauss |
| - | Kelly D. Foote |
| - | Lars Timmermann |
| - | Michael S. Okun |
| - | Patricia Limousin |
| - | Thomas Foltynie |
| - | Veerle Visser-Vandewalle |
| - | Yasin Temel |

Demographic characteristics of the panellists are displayed in Table 4. Briefly, most of them were male (81%), between 50 to 59 years old (47.6%) and highly experienced (95.2% and 85.7% with >10 years of experience in, respectively, neurostimulation field and DBS clinical trials).

**Table 4.** Demographic and academic information for the Delphi Panel members.

|  | *Steering*<br>*Committee*<br>*(n=6)* | *Expert*<br>*Panel*<br>*(n=15)* |
| --- | --- | --- |
| ***Gender*** |  |  |
| Female | 0 | 4 |
| Male | 6 | 11 |
| Prefer not to say | 0 | 0 |
| ***Age (yr)*** |  |  |
| 25-30 | 1 | 0 |
| 31-39 | 2 | 0 |
| 40-49 | 1 | 3 |
| 50-59 | 1 | 9 |
| 60-69 | 1 | 3 |
| Prefer not to say | 0 | 0 |
| ***Highest academic degree*** |  |  |
| Bachelor's Degree | 1 | 0 |
| Master's Degree | 0 | 0 |
| Doctor of Medicine (MD) | 0 | 6 |
| Doctor of Philosophy (PhD) | 5 | 9 |
| Other |  |  |
| ***Country of residence/work*** |  |  |
| Italy | 5 | 0 |
| UK | 0 | 2 |
| Germany | 0 | 6 |
| France | 0 | 1 |
| Canada | 0 | 3 |
| Netherlands | 0 | 1 |
| USA | 1 | 2 |
| ***Primary place of work \**** |  |  |
| Private Company | 1 | 0 |
| Hospital | 2 | 9 |
| University | 3 | 11 |
| Research Institute (public) | 1 | 2 |
| Research Institute (Independent) | 0 | 0 |
| ***Experience in neurostimulation field (yr)*** |  |  |
| ≤5 | 1 | 0 |
| 6-10 | 0 | 0 |
| >10 | 5 | 15 |
| ***field(s) of research (besides neurostimulation) \**** |  |  |
| Biomedical Engineering | 1 | 0 |
| Cognitive Science | 0 | 1 |
| Computational Modelling | 1 | 0 |
| Epidemiology | 0 | 0 |
| Neurology | 4 | 11 |
| Neuroscience | 2 | 8 |
| Neurosurgery | 1 | 6 |
| Pharmacology | 0 | 1 |
| Psychiatry | 0 | 0 |
| Psychology | 0 | 0 |
| Neurorehabilitation | 4 | 3 |
| Other (Systems Neuroscience, EEG, MEG) | 0 | 1 |
| ***Experience in DBS clinical trials (yr)*** |  |  |
| ≤5 | 2 | 0 |
| 6-10 | 1 | 0 |
| >10 | 3 | 15 |
| ***Experience in neurorehabilitation clinical trials (yr)*** |  |  |
| ≤5 | 2 | 6 |
| 6-10 | 2 | 1 |
| >10 | 2 | 8 |

As for the 11 general considerations on PT (Table 5, fig. 2,3,4), the first round led to no consensus for any of the statements; in the second round, the consensus was reached in three statements; finally, in the third round, consensus was reached in four additional statements. In the second round, panellists strongly agreed that PT might help improving motor symptoms (Statement 1) and quality of life (Statement 4) of PwPD undergoing STN-DBS, recommending physical therapists to be part of the multidisciplinary équipe taking care of the patients (Statement 11) (for all, 89% strongly agreed, median ± IQR: 5 ± 0). After the third round, panellists strongly agreed on the need to prescribe PT to PwPD implanted with STN-DBS as soon as the clinical conditions are stable (Statement 8 - 94% strongly agreed, median ± IQR: 5 ± 0) and to chronically-implanted patients (Statement 9 - 88% strongly agreed, median ± IQR: 5 ± 0), because it might help maximizing effects of stimulation (Statement 5 - 88% strongly agreed, median ± IQR: 5 ± 0). Lastly, they suggested PT be prescribed in treatment guidelines as complementary treatment for patients with PwPD treated with STN-DBS (Statement 10 - 88% strongly agreed, median ± IQR: 5 ± 0).

**Fig.2.**
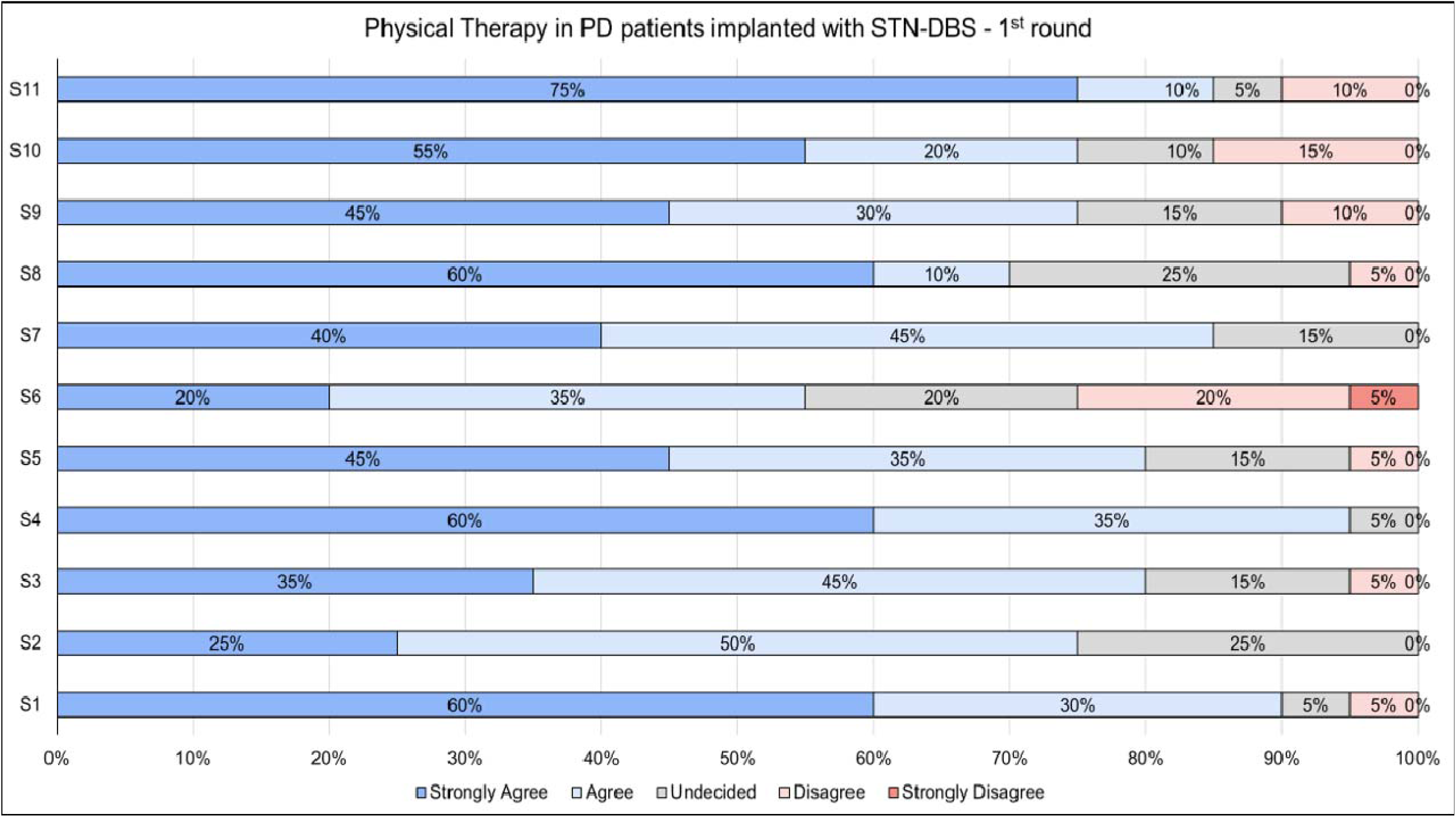
Percentage of agreement for the 11 general considerations on physical therapy after subthalamic nucleus deep brain stimulation in patients with Parkinson’s disease (Statement 1-11) among the Delphi Panel members, as result of the first round. No statement reached a consensus (i.e., >80% of the responses fell in the same response label). PD = Parkinson’s disease; STN-DBS = subthalamic nucleus deep brain stimulation; S = statement.

**Fig.3.**
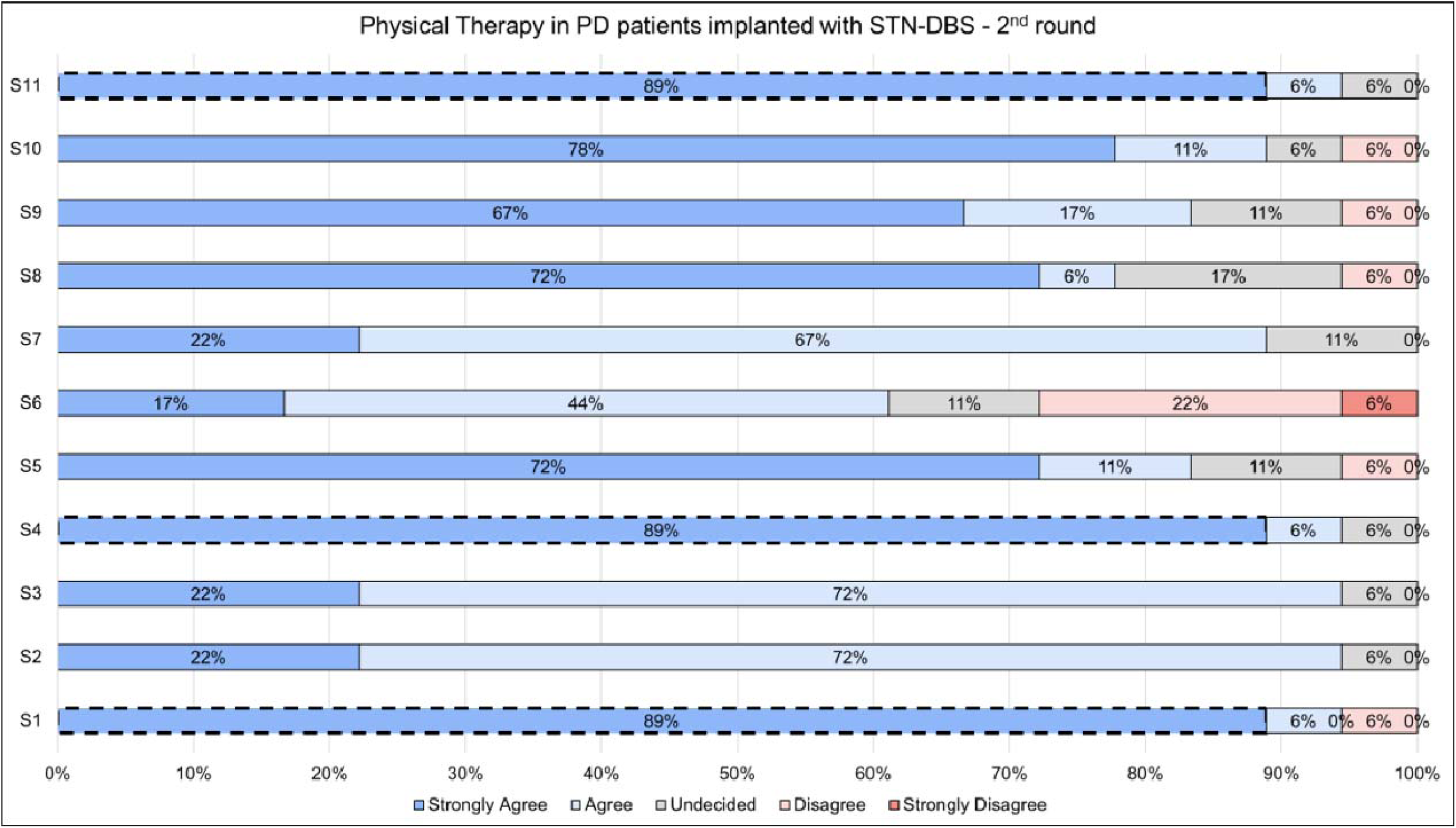
Percentage of agreement for the 11 general considerations on physical therapy after subthalamic nucleus deep brain stimulation in patients with Parkinson’s disease (Statement 1-11) among the Delphi Panel members, as result of the second round. Statement 1, Statement 4 and Statement 11 reached a consensus, i.e., 89% of the responses fell in the response label “strongly agree”. PD = Parkinson’s disease; STN-DBS = subthalamic nucleus deep brain stimulation; S = statement.

**Fig.4.**
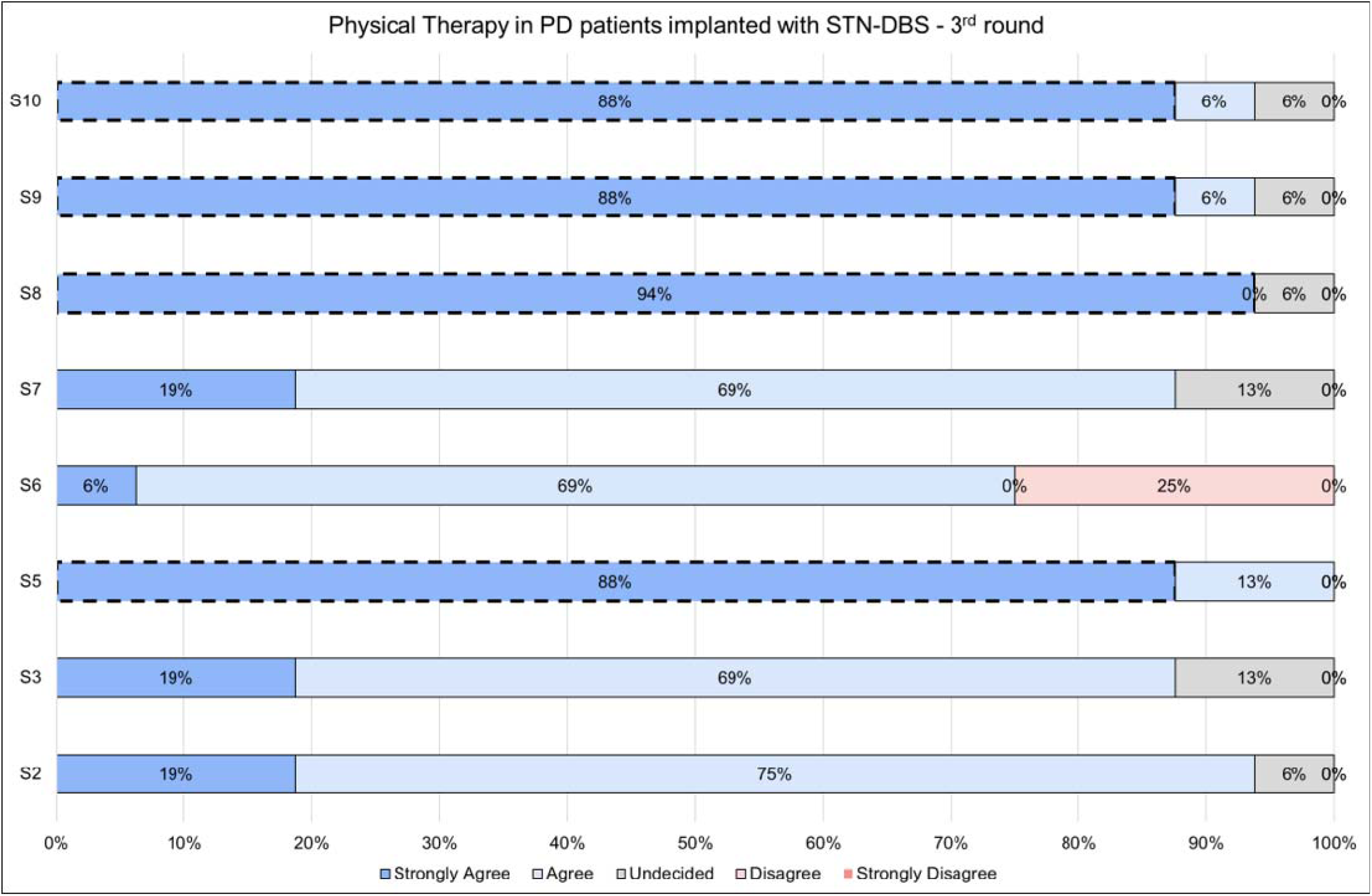
Percentage of agreement for the 11 general considerations on physical therapy after subthalamic nucleus deep brain stimulation in patients with Parkinson’s disease (Statement 1-11) among the Delphi Panel members, as result of the third round. Statement 5, Statement 8, Statement 9 and Statement 10 reached a consensus, i.e., respectively, 88%, 94%, 88% and 88% of the responses fell in the response label “strongly agree”. PD = Parkinson’s disease; STN-DBS = subthalamic nucleus deep brain stimulation; S = statement.

**Table 5.**
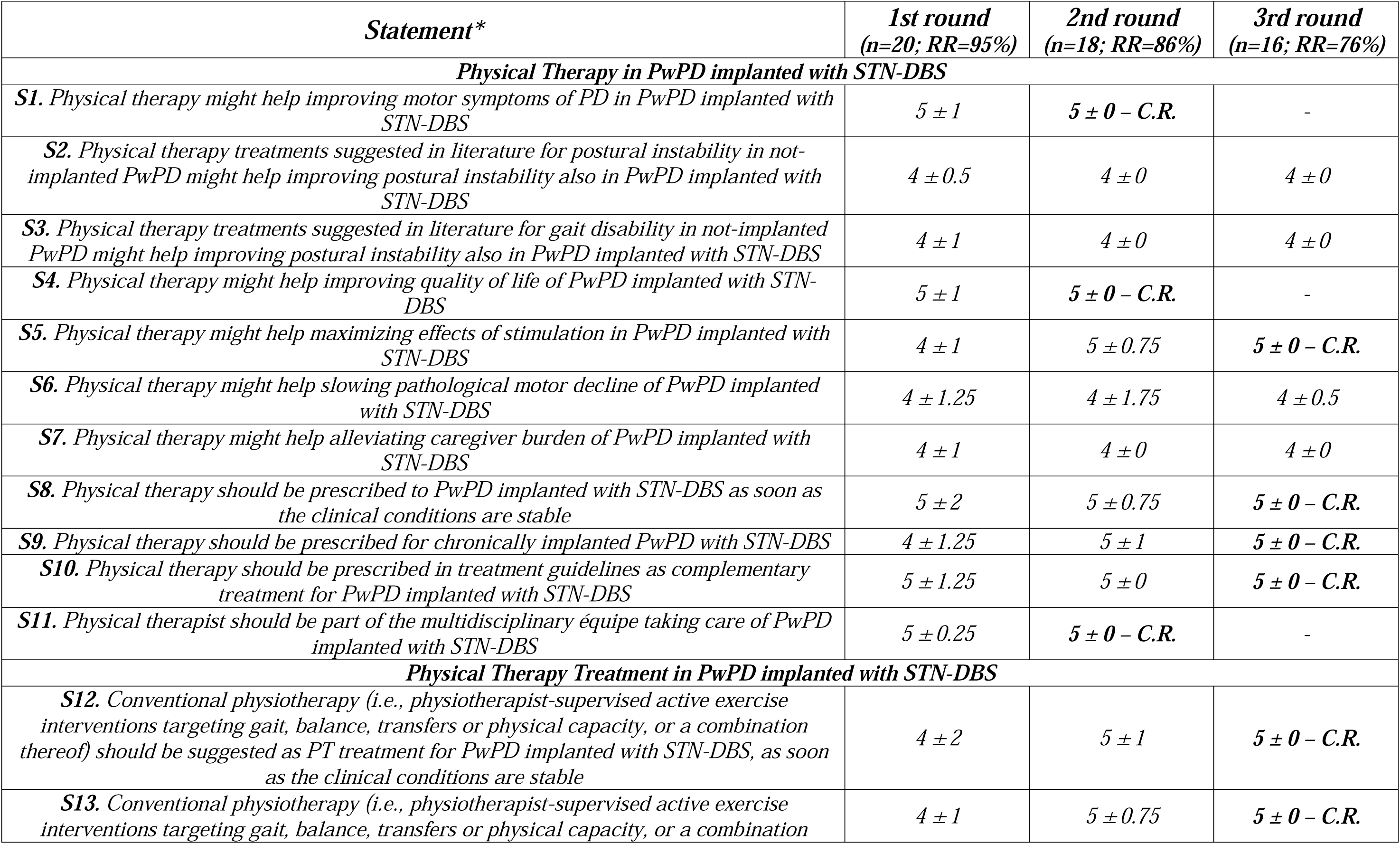
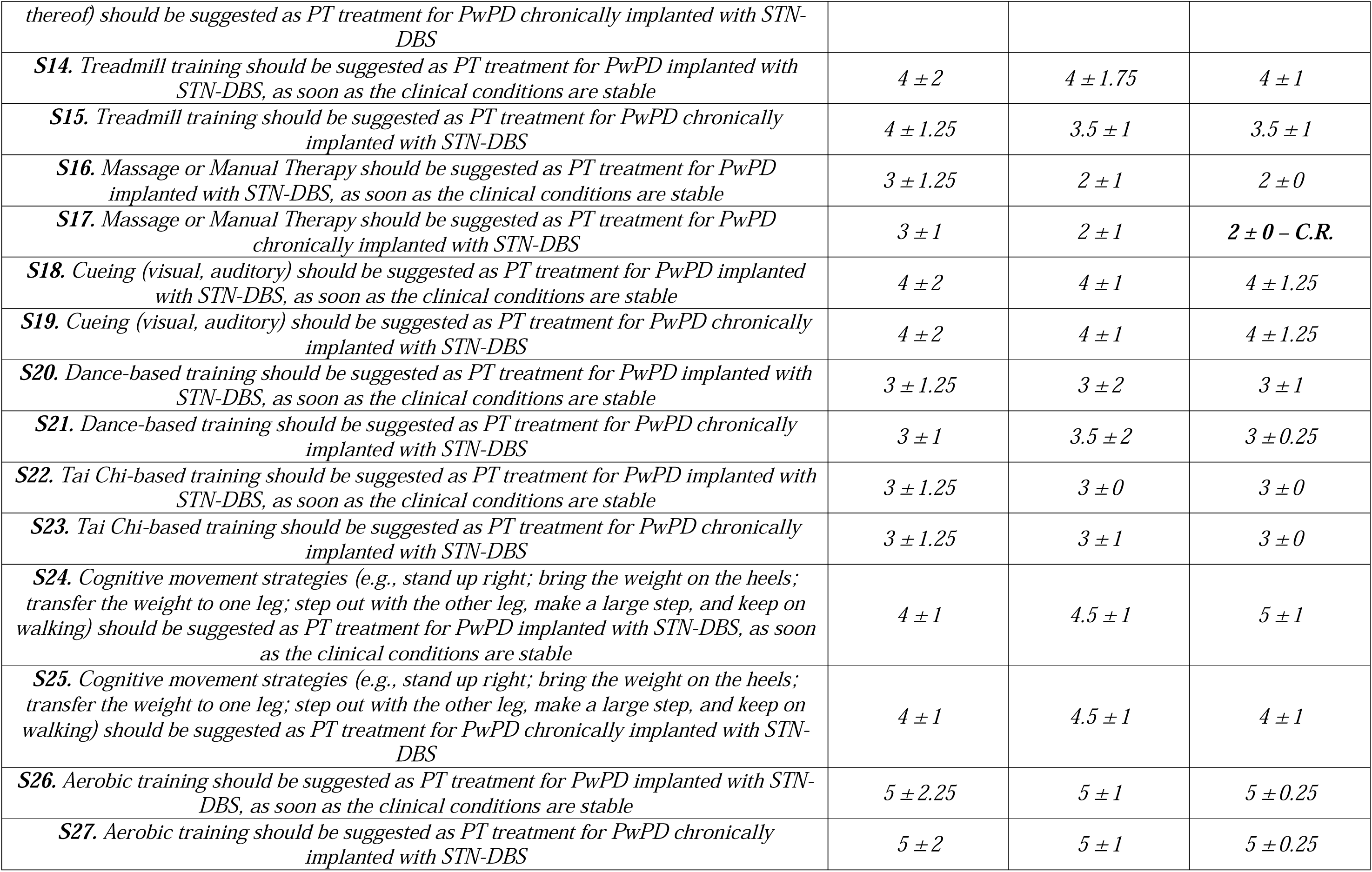
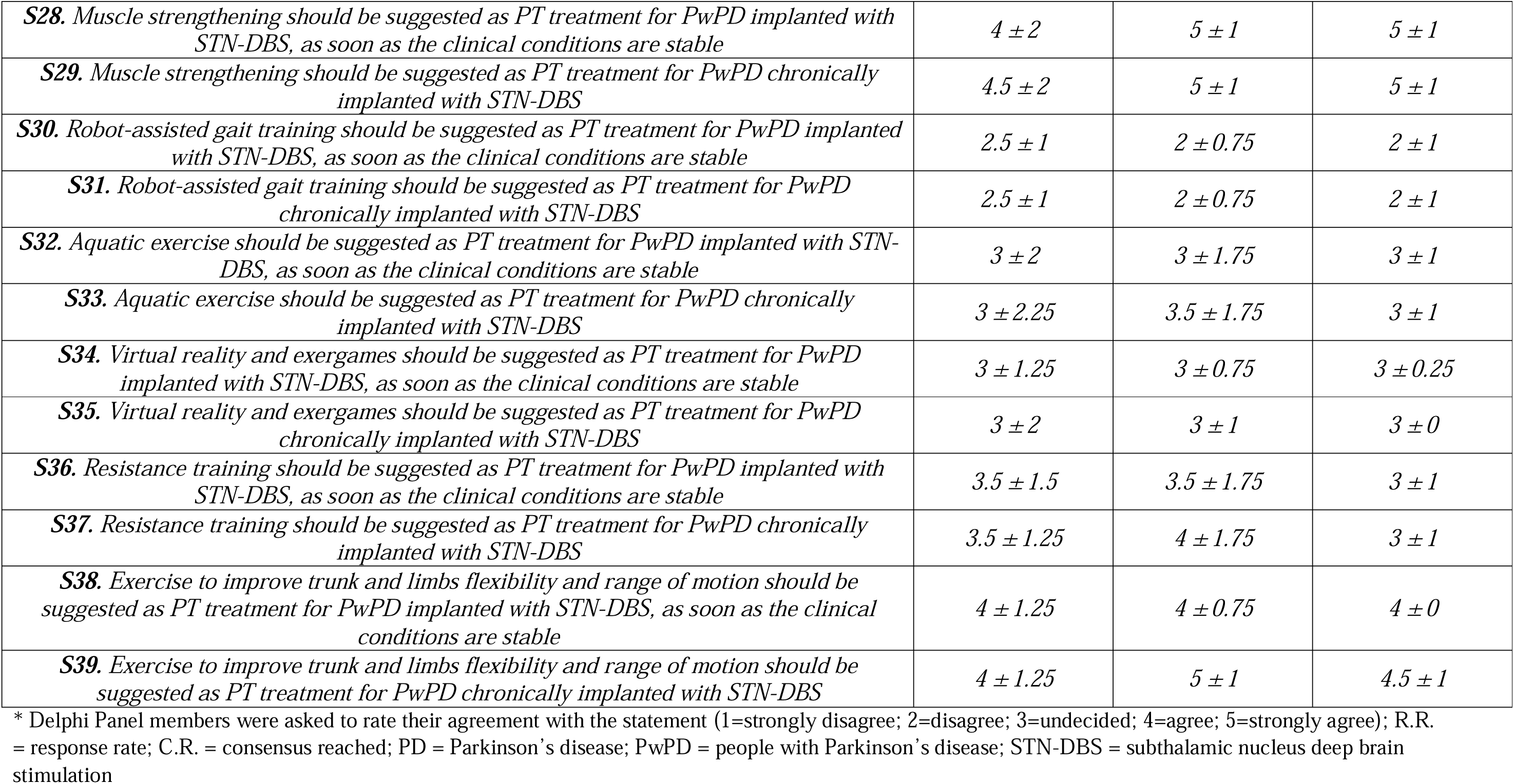
Five-point Likert questionnaire with the results (median ± IQR) for each round.

| *Statement\** | *1st round*<br>(n=20; RR=95%) | *2nd round*<br>(n=18; RR=86%) | *3rd round*<br>(n=16; RR=76%) |
| --- | --- | --- | --- |
| ***Physical Therapy in PwPD implanted with STN-DBS*** |  |  |  |
| *S1. Physical therapy might help improving motor symptoms of PD in PwPD implanted with STN-DBS* | 5 ± 1 | 5 ± 0 – C.R. | - |
| *S2. Physical therapy treatments suggested in literature for postural instability in not-implanted PwPD might help improving postural instability also in PwPD implanted with STN-DBS* | 4 ± 0.5 | 4 ± 0 | 4 ± 0 |
| *S3. Physical therapy treatments suggested in literature for gait disability in not-implanted PwPD might help improving postural instability also in PwPD implanted with STN-DBS* | 4 ± 1 | 4 ± 0 | 4 ± 0 |
| *S4. Physical therapy might help improving quality of life of PwPD implanted with STN-DBS* | 5 ± 1 | 5 ± 0 – C.R. | - |
| *S5. Physical therapy might help maximizing effects of stimulation in PwPD implanted with STN-DBS* | 4 ± 1 | 5 ± 0.75 | 5 ± 0 – C.R. |
| *S6. Physical therapy might help slowing pathological motor decline of PwPD implanted with STN-DBS* | 4 ± 1.25 | 4 ± 1.75 | 4 ± 0.5 |
| *S7. Physical therapy might help alleviating caregiver burden of PwPD implanted with STN-DBS* | 4 ± 1 | 4 ± 0 | 4 ± 0 |
| *S8. Physical therapy should be prescribed to PwPD implanted with STN-DBS as soon as the clinical conditions are stable* | 5 ± 2 | 5 ± 0.75 | 5 ± 0 – C.R. |
| *S9. Physical therapy should be prescribed for chronically implanted PwPD with STN-DBS* | 4 ± 1.25 | 5 ± 1 | 5 ± 0 – C.R. |
| *S10. Physical therapy should be prescribed in treatment guidelines as complementary treatment for PwPD implanted with STN-DBS* | 5 ± 1.25 | 5 ± 0 | 5 ± 0 – C.R. |
| *S11. Physical therapist should be part of the multidisciplinary équipe taking care of PwPD implanted with STN-DBS* | 5 ± 0.25 | 5 ± 0 – C.R. | - |
| ***Physical Therapy Treatment in PwPD implanted with STN-DBS*** |  |  |  |
| *S12. Conventional physiotherapy (i.e., physiotherapist-supervised active exercise interventions targeting gait, balance, transfers or physical capacity, or a combination thereof) should be suggested as PT treatment for PwPD implanted with STN-DBS, as soon as the clinical conditions are stable* | 4 ± 2 | 5 ± 1 | 5 ± 0 – C.R. |
| *S13. Conventional physiotherapy (i.e., physiotherapist-supervised active exercise interventions targeting gait, balance, transfers or physical capacity, or a combination* | 4 ± 1 | 5 ± 0.75 | 5 ± 0 – C.R. |
| *thereof) should be suggested as PT treatment for PwPD chronically implanted with STN-DBS* |  |  |  |
| **S14.** Treadmill training should be suggested as PT treatment for PwPD implanted with STN-DBS, as soon as the clinical conditions are stable | $4 \pm 2$ | $4 \pm 1.75$ | $4 \pm 1$ |
| **S15.** Treadmill training should be suggested as PT treatment for PwPD chronically implanted with STN-DBS | $4 \pm 1.25$ | $3.5 \pm 1$ | $3.5 \pm 1$ |
| **S16.** Massage or Manual Therapy should be suggested as PT treatment for PwPD implanted with STN-DBS, as soon as the clinical conditions are stable | $3 \pm 1.25$ | $2 \pm 1$ | $2 \pm 0$ |
| **S17.** Massage or Manual Therapy should be suggested as PT treatment for PwPD chronically implanted with STN-DBS | $3 \pm 1$ | $2 \pm 1$ | **$2 \pm 0 - C.R.$** |
| **S18.** Cueing (visual, auditory) should be suggested as PT treatment for PwPD implanted with STN-DBS, as soon as the clinical conditions are stable | $4 \pm 2$ | $4 \pm 1$ | $4 \pm 1.25$ |
| **S19.** Cueing (visual, auditory) should be suggested as PT treatment for PwPD chronically implanted with STN-DBS | $4 \pm 2$ | $4 \pm 1$ | $4 \pm 1.25$ |
| **S20.** Dance-based training should be suggested as PT treatment for PwPD implanted with STN-DBS, as soon as the clinical conditions are stable | $3 \pm 1.25$ | $3 \pm 2$ | $3 \pm 1$ |
| **S21.** Dance-based training should be suggested as PT treatment for PwPD chronically implanted with STN-DBS | $3 \pm 1$ | $3.5 \pm 2$ | $3 \pm 0.25$ |
| **S22.** Tai Chi-based training should be suggested as PT treatment for PwPD implanted with STN-DBS, as soon as the clinical conditions are stable | $3 \pm 1.25$ | $3 \pm 0$ | $3 \pm 0$ |
| **S23.** Tai Chi-based training should be suggested as PT treatment for PwPD chronically implanted with STN-DBS | $3 \pm 1.25$ | $3 \pm 1$ | $3 \pm 0$ |
| **S24.** Cognitive movement strategies (e.g., stand up right; bring the weight on the heels; transfer the weight to one leg; step out with the other leg, make a large step, and keep on walking) should be suggested as PT treatment for PwPD implanted with STN-DBS, as soon as the clinical conditions are stable | $4 \pm 1$ | $4.5 \pm 1$ | $5 \pm 1$ |
| **S25.** Cognitive movement strategies (e.g., stand up right; bring the weight on the heels; transfer the weight to one leg; step out with the other leg, make a large step, and keep on walking) should be suggested as PT treatment for PwPD chronically implanted with STN-DBS | $4 \pm 1$ | $4.5 \pm 1$ | $4 \pm 1$ |
| **S26.** Aerobic training should be suggested as PT treatment for PwPD implanted with STN-DBS, as soon as the clinical conditions are stable | $5 \pm 2.25$ | $5 \pm 1$ | $5 \pm 0.25$ |
| **S27.** Aerobic training should be suggested as PT treatment for PwPD chronically implanted with STN-DBS | $5 \pm 2$ | $5 \pm 1$ | $5 \pm 0.25$ |
| **S28.** Muscle strengthening should be suggested as PT treatment for PwPD implanted with STN-DBS, as soon as the clinical conditions are stable | $4 \pm 2$ | $5 \pm 1$ | $5 \pm 1$ |
| **S29.** Muscle strengthening should be suggested as PT treatment for PwPD chronically implanted with STN-DBS | $4.5 \pm 2$ | $5 \pm 1$ | $5 \pm 1$ |
| **S30.** Robot-assisted gait training should be suggested as PT treatment for PwPD implanted with STN-DBS, as soon as the clinical conditions are stable | $2.5 \pm 1$ | $2 \pm 0.75$ | $2 \pm 1$ |
| **S31.** Robot-assisted gait training should be suggested as PT treatment for PwPD chronically implanted with STN-DBS | $2.5 \pm 1$ | $2 \pm 0.75$ | $2 \pm 1$ |
| **S32.** Aquatic exercise should be suggested as PT treatment for PwPD implanted with STN-DBS, as soon as the clinical conditions are stable | $3 \pm 2$ | $3 \pm 1.75$ | $3 \pm 1$ |
| **S33.** Aquatic exercise should be suggested as PT treatment for PwPD chronically implanted with STN-DBS | $3 \pm 2.25$ | $3.5 \pm 1.75$ | $3 \pm 1$ |
| **S34.** Virtual reality and exergames should be suggested as PT treatment for PwPD implanted with STN-DBS, as soon as the clinical conditions are stable | $3 \pm 1.25$ | $3 \pm 0.75$ | $3 \pm 0.25$ |
| **S35.** Virtual reality and exergames should be suggested as PT treatment for PwPD chronically implanted with STN-DBS | $3 \pm 2$ | $3 \pm 1$ | $3 \pm 0$ |
| **S36.** Resistance training should be suggested as PT treatment for PwPD implanted with STN-DBS, as soon as the clinical conditions are stable | $3.5 \pm 1.5$ | $3.5 \pm 1.75$ | $3 \pm 1$ |
| **S37.** Resistance training should be suggested as PT treatment for PwPD chronically implanted with STN-DBS | $3.5 \pm 1.25$ | $4 \pm 1.75$ | $3 \pm 1$ |
| **S38.** Exercise to improve trunk and limbs flexibility and range of motion should be suggested as PT treatment for PwPD implanted with STN-DBS, as soon as the clinical conditions are stable | $4 \pm 1.25$ | $4 \pm 0.75$ | $4 \pm 0$ |
| **S39.** Exercise to improve trunk and limbs flexibility and range of motion should be suggested as PT treatment for PwPD chronically implanted with STN-DBS | $4 \pm 1.25$ | $5 \pm 1$ | $4.5 \pm 1$ |

\* Delphi Panel members were asked to rate their agreement with the statement (1=strongly disagree; 2=disagree; 3=undecided; 4=agree; 5=strongly agree); R.R. = response rate; C.R. = consensus reached; PD = Parkinson's disease; PwPD = people with Parkinson's disease; STN-DBS = subthalamic nucleus deep brain stimulation

As for the 28 statements on PT treatments (Table 5, fig. 5,6,7), no consensus was reached after first and second round. After the third round, consensus was reached in three statements. Indeed, the panellists agreed on the prescription of conventional PT (i.e., physiotherapist-supervised active exercise interventions targeting gait, balance, transfers or physical capacity, or a combination thereof) as soon as the clinical conditions of the implanted patients are stable (Statement 12 - 81% strongly agreed, median ± IQR: 5 ± 0) and in chronically-implanted patients (Statement 13 - 81% strongly agreed, median ± IQR: 5 ± 0). Also, massage or manual therapy was discouraged as treatment for chronically implanted patients (Statement 17 - 81% disagreed, median ± IQR: 2 ± 0).

**Fig.5.**
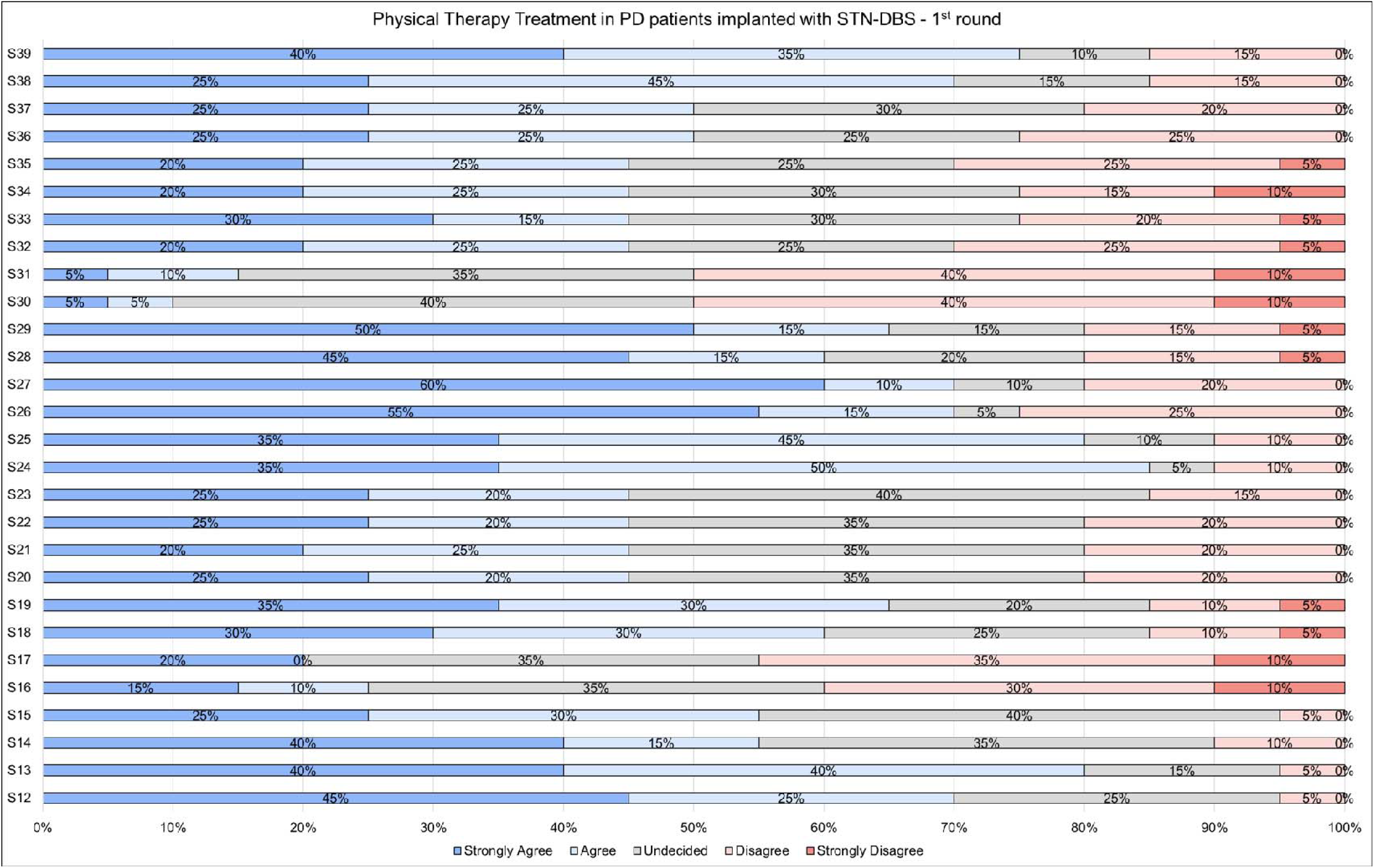
Percentage of agreement for the 28 statements on physical therapy treatments after subthalamic nucleus deep brain stimulation in patients with Parkinson’s disease (Statement 12-39) among the Delphi Panel members, as result of the first round. No statement reached a consensus (i.e., >80% of the responses fell in the same response label). PD = Parkinson’s disease; STN-DBS = subthalamic nucleus deep brain stimulation; S = statement.

**Fig.6.**
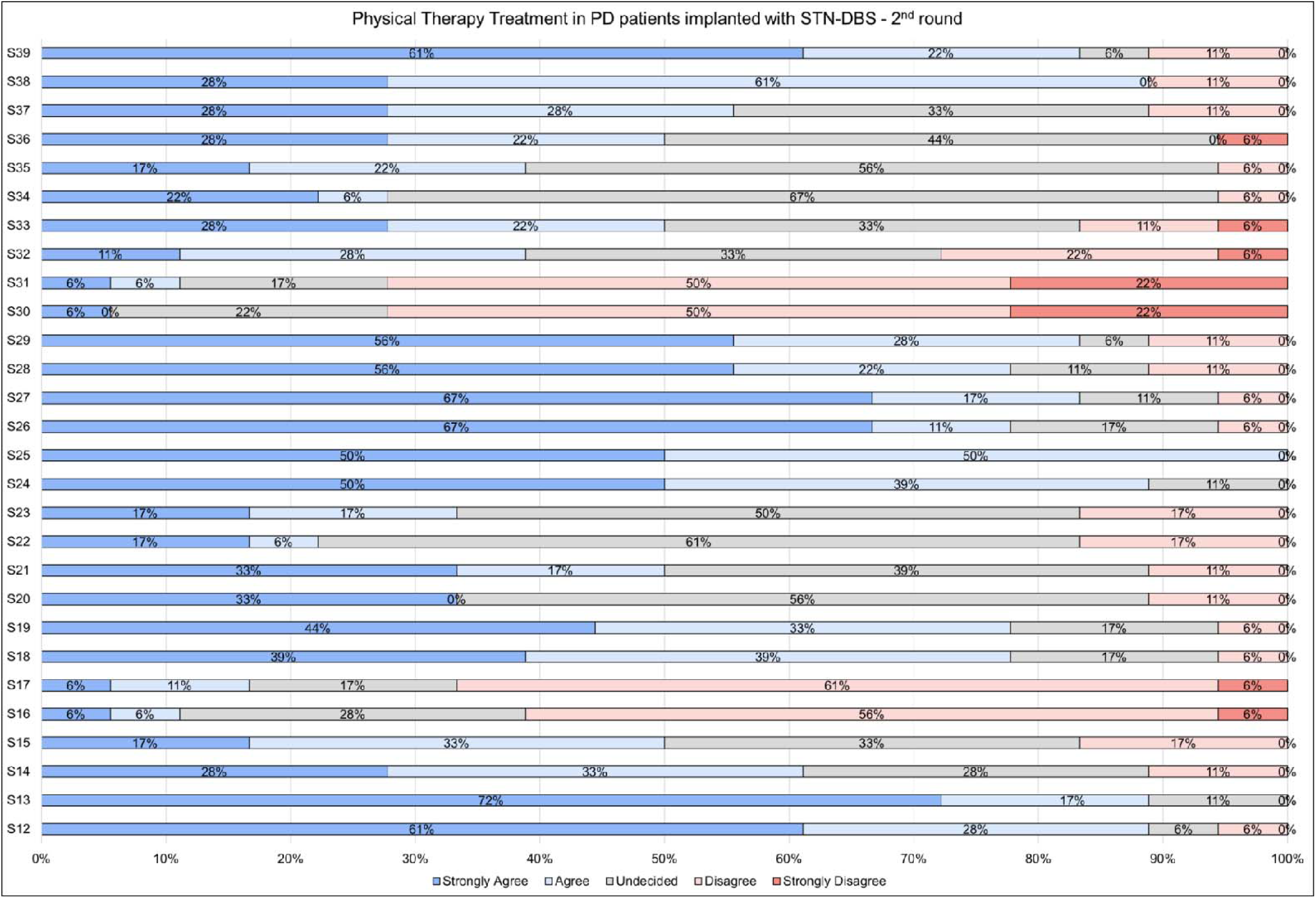
Percentage of agreement for the 28 statements on physical therapy treatments after subthalamic nucleus deep brain stimulation in patients with Parkinson’s disease (Statement 12-39) among the Delphi Panel members, as result of the second round. No statement reached a consensus (i.e., >80% of the responses fell in the same response label). PD = Parkinson’s disease; STN-DBS = subthalamic nucleus deep brain stimulation; S = statement.

**Fig.7.**
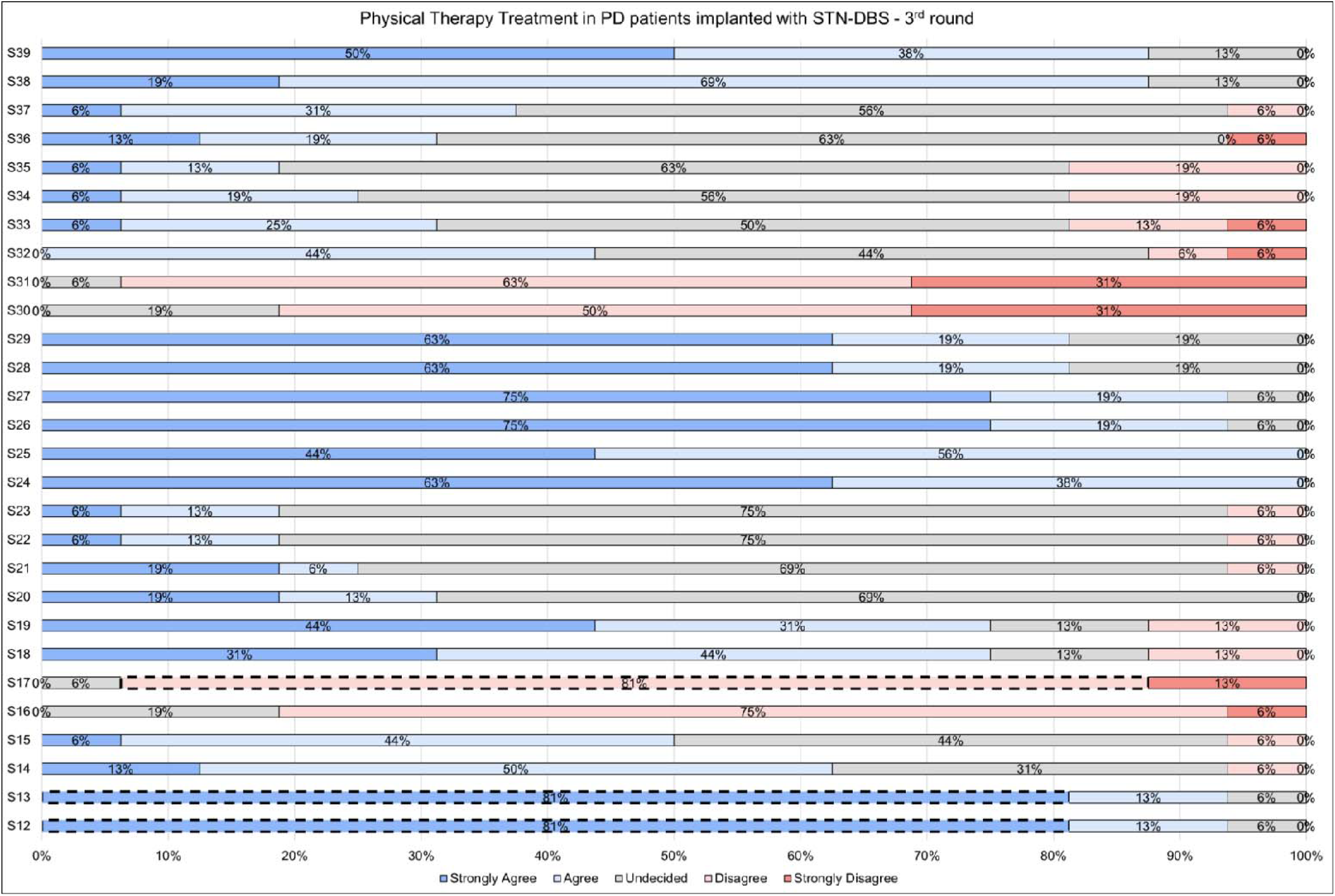
Percentage of agreement for the 28 statements on physical therapy treatments after subthalamic nucleus deep brain stimulation in patients with Parkinson’s disease (Statement 12-39) among the Delphi Panel members, as result of the third round. Statement 12 and Statement 13 reached a consensus, i.e., for both, 81% of the responses fell in the response label “strongly agree”. Statement 17 reached a consensus, i.e., 81% of the responses fell in the response label “disagree”. PD = Parkinson’s disease; STN-DBS = subthalamic nucleus deep brain stimulation; S = statemen

## 4. DISCUSSION

PT is a well-established intervention for psychomotor and functional improvement in the PD general population, with a positive influence on quality of life and independence. However, only very preliminary results are available for PwPD with STN-DBS, even though the combination of DBS and PT may further improve neuromodulation therapy outcomes. The results of our Delphi panel discussion, based on the available knowledge that we summarized in the systematic scoping review here presented, suggest that the scientific community should turn the attention to the lack of knowledge on PT in DBS patients, which represents an actual clinical unmet need for patients, and a limitation for physical therapists in clinical settings.

Our 21 panellists reached a full consensus for 7 out of the 11 statements on the role of PT for PwPD treated with STN-DBS, but only for 3 out of 28 statements on the PT treatments. This is suggestive of the general agreement and acknowledgement on the potential rationale that supports PT intervention in this population, but with clear limitations on the actual protocol of treatment as well as the general need of further research.

### 4.1. PT or no PT?

Considering all the intrinsic caveats and the methodological limitations proper of our systematic scoping review, it might be only qualitatively argued that PT for PwPD treated with STN-DBS could improve dynamic and static balance (61,62,64,67,68,71), gait performance (61–67) and posture (66) (e.g., camptocormia(69)), ultimately leading to significant decrease of daily number of falls (66) and the fear of falling (64), with an increase in motor performances (60,62,64,67,68,71), functional independence (60,67,71), and quality of life (68). Therefore, our expert consensus is of high importance to establish whether PT should be potentially beneficial for PwPD treated with DBS.

Experts agreed that PT might improve motor symptoms and quality of life, maximizing the effects of electrical stimulation. Despite STN-DBS has demonstrated to be highly effective at controlling motor symptoms in PwPD (5), still some clinical issues remain open. After an initial improvement following STN-DBS (73–75) (but see (76)), postural instability in PwPD was reported to worsen 6 months after surgery (77), reaching poorer results than before surgery after 2 years (78). Some findings even suggest no significant improvement in trunk rigidity (6). Similarly, STN-DBS initially improves gait difficulties, but these deficits get worse than before surgery after 2 years (78). Also, gait spatial (e.g., stride length) but not temporal (stride-to-stride variability) parameters were improved after surgery (79) and increased episodes of FOG were reported (80,81), with poorer gait performance on clinical (compared to off stimulation) (81) and subjective assessment (82). Taken these conditions together, they might induce physical inactivity after STN-DBS surgery, increase of falls (83), and secondary complications (84). Interestingly, some authors claimed that stimulation-resistant motor features of PD are often not formally reported (1), either because their relationship with DBS is not recognized or because clinicians fail to routinely document them (85). On the other hand, solid scientific knowledge confirms that PT maximizes independence, well=:Jbeing, and quality of life (9,10), besides improving motor (such as postural instability (13,16,17), gait impairments (18–20) like festination, FOG (9,12,21,22)) and non-motor (e.g., depression, apathy, fatigue (11–15)) PD symptoms. It is reasonable to hypothesize that these evidence in the general PD population would also apply to PwPD implanted with STN-DBS, where exercise and STN-DBS might exert a complimentary, positive effects on PD severity and mobility. This coupled effect has been already shown for exercise and dopaminergic medication on muscle force production, UPDRS III scores, and mobility in PwPD (86). Lastly, both STN-DBS (87) and PT (16) were suggested to stimulate a number of neuroplastic and neuroprotective biochemical events in PwPD. For example, while STN-DBS could preserve nigral dopamine neurons from degeneration (88–91) and raise the level of neurotrophic factors (e.g., BNDF) in the nigrostriatal system and primary motor cortex (92), PT and exercise would enhance neuronal growth, synaptogenesis, neurotrophic factor expression, and neurogenesis (17,23–26). The combination of STN-DBS and PT in PwPD could boost these neurochemical mechanisms and biological pathways, attenuating disease progression and enhancing compensatory neuronal strategies. However, all these assumptions remain to be properly tested. Additionally, it is not clear whether and how PT might affect the frequency with and extent to which STN-DBS parameters might be adjusted over time in PwPD implanted with STN-DBS. It might be reasonable to hypothesize that PwPD implanted with STN-DBS who frequently exercise might require fewer adjustments in their STN-DBS settings over time.

### 4.2. PT prescription

The panel agreed that PT should be prescribed for PwPD implanted with STN-DBS, both in post-acute phase (i.e., as soon as clinical conditions are stable) and in chronic phase. Also, they suggested PT to be included in treatment guidelines, and physical therapists being involved in the multidisciplinary team in charge of the patients. The low-risk nature of PT coupled with the potential benefit for improving motor function and quality of life in PwPD with STN-DBS supports these statements. According to the studies selected in our systematic scoping review, PT in these patients might be considered well tolerated – although the duration of the rehabilitation period might be an obstacle for completion (63). Also, PT appears to be safe, with several studies reporting no intervention-related adverse effects (64,65). For example, Bestaven et al. (66) reported that, despite initial doubts and apprehension, all the subjects enrolled accepted and completed the PT protocol. Also, current recommendations allow return to exercise within weeks following surgery (93), therefore it appears that PT should be considered a not-harmful intervention for PwPD with STN-DBS, even more so because PT is commonly a supervised treatment. Physical therapists, indeed, according to their training and specific skills, could contribute to take care of the patients after implantation surgery (e.g., in the management of complications after surgery (94,95) or during the adaptation of the stimulation parameters (94,95)) or in the chronic phase (e.g., modifying pathological movement patterns (94) or teaching the patients to adapt motor strategies and relevant activities of daily living to the new conditions (94)). Besides technical aspects of intervention, PT treatment characteristically requires multiple sessions (several times a week) for quite long periods (weeks to months) (96) – a time where patient-therapist relationship can be developed for explanations or counselling (97). This could represent an occasion to increase cooperation and motivation from patients and caregivers, which is fundamental to achieve a good outcome after DBS (98).

### 4.3. PT protocols

Our systematic scoping review revealed that the current studies applied different treatments in terms of types of exercise and protocol (frequency, duration), which were assessed through a variety of outcomes only-partially comparable. Despite this very limited scientific knowledge, panellists agreed that conventional PT (i.e., physiotherapist-supervised active exercise interventions targeting gait, balance, transfers, or physical capacity) should be prescribed as a rehabilitative treatment for PwPD implanted with STN-DBS, regardless the time from the surgery. This is probably because most of the limited knowledge on PT treatments in PwPD undergoing STN-DBS is related to “conventional PT”-like interventions (61,62,65–67,69,71), as shown by our systematic scoping review. The results suggest a positive effect on motor and functional PD symptoms (61,62,65–67,69,71). A number of findings suggest similar effects for general PD population (15), although without superiority over other types of treatment (96). For example, several studies suggest that multifactorial conventional PT interventions including muscle strengthening, increasing of range of movement, balance training and gait training have a positive effect on balance dysfunction and postural instability in PwPD (16,99). Also, balance training improves the self-confidence perceived while performing activities of daily life and reduces falls rate (18), while gait training improves FOG, gait speed and step length, even for months after the treatment. (19,22,100). It is plausible to speculate that PwPD with STN-DBS implants might benefit from the same evidence seen in the general PD population. However, this still needs to be thoroughly examined. Indeed, it is not clear, at this stage of knowledge, which PT treatments should be included as standard of care following STN-DBS, nor their characteristics (e.g., frequency, intensity) or the factors to be considered for a treatment stratification (e.g., time from surgery, characteristics of the stimulation, effects of the stimulation)(4) – a concerted effort is needed to explore PT interventions, and solidify PT as part of the post-surgical standard of care for PwPD with STN-DBS.

On the other side, panellists agreed that massage or manual therapy should not be applied in chronically implanted patients. While no evidence is currently available to support or discourage these treatment approaches in PwPD treated with STN-DBS, a systematic review suggests that the evidence on the effectiveness of massage therapy in the general PD population is limited and conflicting in some cases due to methodological concerns (101). The European Physiotherapy Guideline for Parkinson’s disease released a weak recommendation for using massage or manual therapy to reduce pain and muscular spasms, but highlighted the need to always combine it with other types of intervention as no evidence support their use to improve physical and functional performance (32). More studies and efforts are needed to elucidate the role of this passive techniques in PwPD with STN-DBS implantation, even more when their role is unclear in general PD population.

### 4.4. Differences to be considered

The main domain of rehabilitation is the correct management of the complex interaction between functioning, health condition, personal factors and environment to improve quality of life and reduce disability (94). This is of particular importance in PwPD treated with STN-DBS, because DBS surgery typically comes after several years of pathology, during which the patient has modified his/her body image (102), motor behaviour (103), environment and lifestyle (104,105) as consequence of or to compensate the progression of the disease. The stimulation induces a sudden and rapid neurophysiological and clinical change, although for the better, that requires the active and participatory role of the patient to readapt motor and functional strategies. This is essential to fully exploit the potentiality of improvement given by DBS – motor readaptation which can be achieved through PT. For example, pathological movement patterns typical of gait in PD (e.g., increase activity of leg flexors and decreased activity of leg extensors in stance phases of gait (106)), which lead to instability, has to be gradually adapted to the better mobility achieved by STN-DBS (94). Therefore, PwPD undergoing STN-DBS will reach a clinical stage where rehabilitation strategies must be implemented in order to support the readaptation of motor and functional strategies following the improvement gained with DBS.

It was proposed that general principles of motor rehabilitation for PD are applicable to those patients undergoing DBS (94). These include the personalization of motor strategies (i.e., awareness of multiple motor strategies and the selection of the appropriate one) and application of the principles of motor learning (e.g., repetition, shaping, massed practice, task-specific training) (107). However, some critical differences from general PD population that PT programs must consider might be identified:

I. Pre-surgery characteristics of the patients. According to the Core Assessment Program for Surgical Interventional Therapies in PD (CAPSIT-PD) (108), STN-DBS surgery candidate should meet several inclusion criteria before undergoing the stimulation. Commonly selected patients have a relatively young onset of PD and an age below 70 years at the time of surgery (98), but they are on relatively advanced stages of the disease, with significant motor problems and a disease duration of 12-15 years (109). Nonetheless, their cognitive (i.e., absence of dementia (109)) and psychiatric (i.e., absence of unstable psychiatric condition (98)) profile are substantially satisfactory. Also, they have a good responsiveness to levodopa (i.e., motor symptoms improve at least by 30% after intake) (108,109), and levodopa-unresponsive symptoms (e.g., gait and balance issues, dysphagia, dysarthria) are not severe (98). All these characteristics must be considered while studying the role of PT in PwPD with STN-DBS, and PT programs tailored.
II. Actual clinical characteristics of the patients. Since STN-DBS can improve only some motor symptoms, a specific clinical pattern was proposed for PwPD with STN-DBS, where tremor, rigidity, bradykinesia, on–off fluctuations and dyskinesias are well-controlled, but gait, stability, postural abnormalities (but also speech and cognition) impairments are still present (5,6). Therefore, these should be the primary targets of PT interventions.
III. Presence of the hardware. DBS implant requires that electrodes, inserted through a small opening in the skull and implanted into the brain, are connected (through extension wire passing under the skin of head, neck, and shoulder) to a battery source which is implanted under the skin in the upper chest (110). A systematic review of hardware-related complications of DBS reported that lead migration or dislocation and fracture or failure of some parts of the DBS system are among the most common complications after DBS surgery (111). Some studies estimated migration or misplacement of the leads and lead fractures among, respectively, 0%-19% and 0%-15% (83,112,113). Therefore, although the risks of issues related to the DBS hardware components are low, and PT programs studied appear to be safe, a more intensive research program must consider hardware presence and frailty when developing or optimizing PT strategies. Besides these aspects, the use of any physical forces (e.g., magnetic fields) that could interfere with DBS components should be avoided to guarantee the correct functioning of stimulation.
IV. Interaction between stimulation and PT: in the light of the opportunities given by advanced DBS technologies (2), such as adaptive DBS (114), it is likely that the patient might need specific DBS programming while undergoing PT sessions, in order to boost their performance and to optimize benefits. This should be a further research topic to be considered while physiotherapists and DBS experts will interact to develop effective and personalized rehabilitation programs.

### 4.5. Limitations

The consensus among experts reached through the Delphi method offers only the weakest degree of support for inferring causal relationships. (115) Therefore, the panel conclusions should not be viewed as a replacement for clinical judgment or original research. Rather, our results are relevant mostly in terms of future research directions, which will foster the development of the field of rehabilitation after STN-DBS in PwPD, because they are based on the collective expertise of a panel of experts who can draw on both their personal experience and scientific knowledge. Indeed, more discussion and empirical evidence are needed to support the feasibility of our results, especially considering that great lack of high-quality evidence which currently characterize these topics.

## 5. CONCLUSION

Despite the limited, low-quality knowledge currently available on the role of PT in PwPD and STN-DBS, panellists agreed that PT may aid in improving motor symptoms and quality of life of this population of patients, probably through a maximization of stimulation effects. They agreed that PT should be prescribed, regardless of the time from the surgery, and that PT should be included in management guidelines, with physical therapists integrated in the multidisciplinary team taking care of the patients. Regarding the type of PT treatment, panellists suggested that conventional PT (but not massage or manual therapy) should be considered. In conclusion, the results of this Delphi consensus represent a call to both the rehabilitation and the DBS community to start working and interacting to deepen this field of research, which for many years has been relegated to the personal expertise of physical therapists despite the increasing number of PwPD implanted with STN-DBS.

## Data Availability

All data produced in the present study are available upon reasonable request to the authors

## Acknowledgment

The authors acknowledge Dr. Briselda Doka, who contributed to early stages of the literature research and data handling.

## REFERENCE

1. Limousin P, Foltynie T. Long-term outcomes of deep brain stimulation in Parkinson disease. Nature Reviews Neurology 2019 15:4. 18 febbraio 2019;15(4):234–42.

2. Guidetti M, Marceglia S, Loh A, Harmsen IE, Meoni S, Foffani G, et al. Clinical perspectives of adaptive deep brain stimulation. Brain stimulation. 1 settembre 2021;14(5):1238–47.

3. Conway ZJ, Silburn PA, Thevathasan W, Maley KO, Naughton GA, Cole MH. Alternate Subthalamic Nucleus Deep Brain Stimulation Parameters to Manage Motor Symptoms of Parkinson’s Disease: Systematic Review and Meta-analysis. Movement Disorders Clinical Practice. 1 gennaio 2019;6(1):17–26.

4. Fasano A, Aquino CC, Krauss JK, Honey CR, Bloem BR. Axial disability and deep brain stimulation in patients with Parkinson disease. Nat Rev Neurol. febbraio 2015;11(2):98–110.

5. Lozano AM, Lipsman N, Bergman H, Brown P, Chabardes S, Chang JW, et al. Deep brain stimulation: current challenges and future directions. Nat Rev Neurol. marzo 2019;15(3):148– 60.

6. Rodriguez-Oroz MC, Moro E, Krack P. Long-term outcomes of surgical therapies for Parkinson’s disease. Mov Disord. dicembre 2012;27(14):1718–28.

7. Rubenis J. A rehabilitational approach to the management of Parkinson’s disease. Parkinsonism & Related Disorders. 1 gennaio 2007;13(SUPPL. 3):S495–7.

8. Playfer JR, Hindle JV. Rehabilitation and the interdisciplinary team. 19 aprile 2018;291–313.

9. Keus SHJ, Bloem BR, Hendriks EJM, Bredero-Cohen AB, Munneke M. Evidence-based analysis of physical therapy in Parkinson’s disease with recommendations for practice and research. Movement Disorders. 15 marzo 2007;22(4):451–60.

10. Osborne JA, Botkin R, Colon-Semenza C, DeAngelis TR, Gallardo OG, Kosakowski H, et al. Physical Therapist Management of Parkinson Disease: A Clinical Practice Guideline From the American Physical Therapy Association. Phys Ther. 1 aprile 2022;102(4):pzab302.

11. de Dreu MJ, van der Wilk ASD, Poppe E, Kwakkel G, van Wegen EEH. Rehabilitation, exercise therapy and music in patients with Parkinson’s disease: a meta-analysis of the effects of music-based movement therapy on walking ability, balance and quality of life. Parkinsonism & related disorders [Internet]. 2012 [citato 12 maggio 2022];18 Suppl 1(SUPPL. 1). Disponibile su: https://pubmed.ncbi.nlm.nih.gov/22166406/

12. Goodwin VA, Richards SH, Taylor RS, Taylor AH, Campbell JL. The effectiveness of exercise interventions for people with Parkinson’s disease: a systematic review and meta-analysis. Movement disorders=:J: official journal of the Movement Disorder Society. 15 aprile 2008;23(5):631–40.

13. Tomlinson CL, Patel S, Meek C, Clarke CE, Stowe R, Shah L, et al. Physiotherapy versus placebo or no intervention in Parkinson’s disease. The Cochrane database of systematic reviews [Internet]. 11 luglio 2012 [citato 12 maggio 2022];(7). Disponibile su: https://pubmed.ncbi.nlm.nih.gov/22786482/

14. Okada Y, Ohtsuka H, Kamata N, Yamamoto S, Sawada M, Nakamura J, et al. Effectiveness of Long-Term Physiotherapy in Parkinson’s Disease: A Systematic Review and Meta-Analysis. Journal of Parkinson’s disease. 2021;11(4):1619–30.

15. Radder DLM, Lígia Silva de Lima A, Domingos J, Keus SHJ, van Nimwegen M, Bloem BR, et al. Physiotherapy in Parkinson’s Disease: A Meta-Analysis of Present Treatment Modalities. Neurorehabilitation and neural repair. 1 ottobre 2020;34(10):871–80.

16. Hirsch MA, Toole T, Maitland CG, Rider RA. The effects of balance training and high-intensity resistance training on persons with idiopathic Parkinson’s disease. Archives of Physical Medicine and Rehabilitation. 1 agosto 2003;84(8):1109–17.

17. Abbruzzese G, Marchese R, Avanzino L, Pelosin E. Rehabilitation for Parkinson’s disease: Current outlook and future challenges. Parkinsonism & related disorders. 1 gennaio 2016;22 Suppl 1:S60–4.

18. Smania N, Corato E, Tinazzi M, Stanzani C, Fiaschi A, Girardi P, et al. Effect of balance training on postural instability in patients with idiopathic parkinsong’s disease. Neurorehabilitation and Neural Repair. 2 novembre 2010;24(9):826–34.

19. Bello O, Sanchez JA, Lopez-Alonso V, Márquez G, Morenilla L, Castro X, et al. The effects of treadmill or overground walking training program on gait in Parkinson’s disease. Gait & Posture. 1 settembre 2013;38(4):590–5.

20. Smania N, Corato E, Tinazzi M, Stanzani C, Fiaschi A, Girardi P, et al. Effect of balance training on postural instability in patients with idiopathic parkinsong’s disease. Neurorehabilitation and Neural Repair. 2 novembre 2010;24(9):826–34.

21. Fox SH, Katzenschlager R, Lim SY, Ravina B, Seppi K, Coelho M, et al. The Movement Disorder Society Evidence-Based Medicine Review Update: Treatments for the motor symptoms of Parkinson’s disease. Movement Disorders. 1 ottobre 2011;26(S3):S2–41.

22. Cosentino C, Baccini M, Putzolu M, Ristori D, Avanzino L, Pelosin E. Effectiveness of Physiotherapy on Freezing of Gait in Parkinson’s Disease: A Systematic Review and Meta-Analyses. Movement disorders=:J: official journal of the Movement Disorder Society. 1 aprile 2020;35(4):523–36.

23. Policastro G, Brunelli M, Tinazzi M, Chiamulera C, Emerich DF, Paolone G. Cytokine-, Neurotrophin-, and Motor Rehabilitation-Induced Plasticity in Parkinson’s Disease. Neural plasticity [Internet]. 2020 [citato 12 maggio 2022];2020. Disponibile su: https://pubmed.ncbi.nlm.nih.gov/33293946/

24. Małczyńska-Sims P, Chalimoniuk M, Sułek A. The Effect of Endurance Training on Brain-Derived Neurotrophic Factor and Inflammatory Markers in Healthy People and Parkinson’s Disease. A Narrative Review. Frontiers in physiology [Internet]. 19 novembre 2020 [citato 12 maggio 2022];11. Disponibile su: https://pubmed.ncbi.nlm.nih.gov/33329027/

25. Torikoshi S, Morizane A, Shimogawa T, Samata B, Miyamoto S, Takahashi J. Exercise Promotes Neurite Extensions from Grafted Dopaminergic Neurons in the Direction of the Dorsolateral Striatum in Parkinson’s Disease Model Rats. Journal of Parkinson’s disease. 2020;10(2):511–21.

26. Xu X, Fu Z, Le W. Exercise and Parkinson’s disease. International review of neurobiology. 1 gennaio 2019;147:45–74.

27. Duncan RP, Earhart GM. Randomized controlled trial of community-based dancing to modify disease progression in Parkinson disease. Neurorehabilitation and neural repair. febbraio 2012;26(2):132–43.

28. Ellis T, Katz DI, White DK, DePiero TJ, Hohler AD, Saint-Hilaire M. Effectiveness of an inpatient multidisciplinary rehabilitation program for people with Parkinson disease. Physical therapy. luglio 2008;88(7):812–9.

29. Li F, Harmer P, Fitzgerald K, Eckstrom E, Stock R, Galver J, et al. Tai chi and postural stability in patients with Parkinson’s disease. The New England journal of medicine. 9 febbraio 2012;366(6):511–9.

30. Prodoehl J, Rafferty MR, David FJ, Poon C, Vaillancourt DE, Comella CL, et al. Two-year exercise program improves physical function in Parkinson’s disease: the PRET-PD randomized clinical trial. Neurorehabilitation and neural repair. 2 marzo 2015;29(2):112–22.

31. Schenkman M, Hall DA, Baron AE, Schwartz RS, Mettler P, Kohrt WM. Exercise for people in early-or mid-stage Parkinson disease: a 16-month randomized controlled trial. Physical therapy. novembre 2012;92(11):1395–410.

32. Keus S, Munneke M, Graziano M, Paltamaa J, Pelosin E, Domingos J, et al. European Physiotherapy Guideline for Parkinson’s Disease Developed with twenty European professional associations. 2014 [citato 13 maggio 2022]; Disponibile su: www.parkinsonnet.info/euguideline

33. Peters MDJ, Godfrey CM, Khalil H, McInerney P, Parker D, Soares CB. Guidance for conducting systematic scoping reviews. JBI Evidence Implementation. settembre 2015;13(3):141.

34. Tricco AC, Lillie E, Zarin W, O’Brien KK, Colquhoun H, Levac D, et al. PRISMA Extension for Scoping Reviews (PRISMA-ScR): Checklist and Explanation. Ann Intern Med. 2 ottobre 2018;169(7):467–73.

35. Arksey H, O’Malley L. Scoping studies: towards a methodological framework. International journal of social research methodology. 2005;8(1):19–32.

36. Pham MT, Rajić A, Greig JD, Sargeant JM, Papadopoulos A, McEwen SA. A scoping review of scoping reviews: advancing the approach and enhancing the consistency. Res Synth Methods. dicembre 2014;5(4):371–85.

37. Colquhoun HL, Levac D, O’Brien KK, Straus S, Tricco AC, Perrier L, et al. Scoping reviews: time for clarity in definition, methods, and reporting. J Clin Epidemiol. dicembre 2014;67(12):1291–4.

38. Daudt HM, van Mossel C, Scott SJ. Enhancing the scoping study methodology: a large, inter-professional team’s experience with Arksey and O’Malley’s framework. BMC Medical Research Methodology. 23 marzo 2013;13(1):48.

39. Downs SH, Black N. The feasibility of creating a checklist for the assessment of the methodological quality both of randomised and non-randomised studies of health care interventions. J Epidemiol Community Health. giugno 1998;52(6):377–84.

40. Price OJ, Sewry N, Schwellnus M, Backer V, Reier-Nilsen T, Bougault V, et al. Prevalence of lower airway dysfunction in athletes: a systematic review and meta-analysis by a subgroup of the IOC consensus group on ‘acute respiratory illness in the athlete’. Br J Sports Med. 1 febbraio 2022;56(4):213–22.

41. Kerlinger FN. Foundations of Behavioral Research New York: Holt. Rinehart & Winston. 1973;

42. Lynn MR, Layman EL, Englebardt SP. Nursing administration research priorities. A national Delphi study. J Nurs Adm. maggio 1998;28(5):7–11.

43. McKenna HP. The Delphi technique: a worthwhile research approach for nursing? J Adv Nurs. giugno 1994;19(6):1221–5.

44. Ludwig B. Predicting the future: Have you considered using the Delphi methodology? Journal of extension. 1997;

45. Turoff M, Hiltz SR. Computer based Delphi processes. Gazing into the oracle: The Delphi method and its application to social policy and public health. 1996;56–85.

46. Ulschak FL. Human resource development: The theory and practice of need assessment. Reston Publishing Company; 1983.

47. Rowe G, Wright G. Expert Opinions in Forecasting: The Role of the Delphi Technique. In: Armstrong JS, curatore. Principles of Forecasting: A Handbook for Researchers and Practitioners [Internet]. Boston, MA: Springer US; 2001 [citato 9 giugno 2023]. p. 125–44. (International Series in Operations Research & Management Science). Disponibile su: 10.1007/978-0-306-47630-3_7

48. Brooks KW. Delphi technique: Expanding applications. North Central Association Quarterly. 1979;53(3):377–85.

49. Custer RL, Scarcella JA, Stewart BR. The modified Delphi technique-A rotational modification. 1999;

50. Cyphert FR, Gant WL. The delphi technique: A case study. Phi Delta Kappan. 1971;52(5):272–3.

51. Ludwig BG. Internationalizing Extension: An exploration of the characteristics evident in a state university Extension system that achieves internationalization. The Ohio State University; 1994.

52. Hsu CC, Sandford BA. The Delphi technique: making sense of consensus. Practical assessment, research, and evaluation. 2007;12(1):10.

53. Hasson F, Keeney S, McKenna H. Research guidelines for the Delphi survey technique. Journal of Advanced Nursing. 2000;32(4):1008–15.

54. Kaplan LM. The use of the Delphi method in organizational communication: A case study. Ohio State University; 1971.

55. Sumsion T. The Delphi Technique: An Adaptive Research Tool. British Journal of Occupational Therapy. 1 aprile 1998;61(4):153–6.

56. Green B, Jones M, Hughes D, Williams A. Applying the Delphi technique in a study of GPs’ information requirements. Health Soc Care Community. maggio 1999;7(3):198–205.

57. Eckman CA. DEVELOPMENT OF AN INSTRUMENT TO EVALUATE INTERCOLLEGIATE ATHLETIC COACHES: A MODIFIED DELPHI STUDY. 1984;

58. Hill KQ, Fowles J. The methodological worth of the Delphi forecasting technique. Technological forecasting and social change. 1975;7(2):179–92.

59. Jacobs JM. Essential assessment criteria for physical education teacher education programs: A Delphi study. West Virginia University; 1996.

60. Cohen DB, Oh MY, Baser SM, Angle C, Whiting A, Birk C, et al. Fast-track programming and rehabilitation model: a novel approach to postoperative deep brain stimulation patient care. Arch Phys Med Rehabil. ottobre 2007;88(10):1320–4.

61. Sato K, Aita N, Hokari Y, Kitahara E, Tani M, Izawa N, et al. Balance and Gait Improvements of Postoperative Rehabilitation in Patients with Parkinson’s Disease Treated with Subthalamic Nucleus Deep Brain Stimulation (STN-DBS). Parkinsons Dis. 2019;2019:7104071.

62. Sato K, Hokari Y, Kitahara E, Izawa N, Hatori K, Honaga K, et al. Short-Term Motor Outcomes in Parkinson’s Disease after Subthalamic Nucleus Deep Brain Stimulation Combined with Post-Operative Rehabilitation: A Pre-Post Comparison Study. Parkinsons Dis. 2022;2022:8448638.

63. Nardo A, Anasetti F, Servello D, Porta M. Quantitative gait analysis in patients with Parkinson treated with deep brain stimulation: the effects of a robotic gait training. NeuroRehabilitation. 2014;35(4):779–88.

64. Naro A, Pignolo L, Sorbera C, Latella D, Billeri L, Manuli A, et al. A Case-Controlled Pilot Study on Rhythmic Auditory Stimulation-Assisted Gait Training and Conventional Physiotherapy in Patients With Parkinson’s Disease Submitted to Deep Brain Stimulation. Front Neurol. 2020;11:794.

65. Luna NMS, Lucareli PRG, Sales VC, Speciali D, Alonso AC, Peterson MD, et al. Treadmill training in Parkinson’s patients after deep brain stimulation: Effects on gait kinematic. NeuroRehabilitation. 1 gennaio 2018;42(2):149–58.

66. Bestaven E, Guillaud E, De Sèze M, Jerome A, Burbaud P, Cazalets JR, et al. Effect of Trunk Muscle Strengthening on Gait Pattern and Falls in Parkinson’s Disease. J Rehabil Med Clin Commun. 2019;2:1000003.

67. Tassorelli C, Buscone S, Sandrini G, Pacchetti C, Furnari A, Zangaglia R, et al. The role of rehabilitation in deep brain stimulation of the subthalamic nucleus for Parkinson’s disease: a pilot study. Parkinsonism Relat Disord. novembre 2009;15(9):675–81.

68. Li H, Liang S, Yu Y, Wang Y, Cheng Y, Yang H, et al. Clinical experience of comprehensive treatment on the balance function of Parkinson’s disease. Medicine (Baltimore). maggio 2020;99(19):e20154.

69. Liang S, Yu Y, Li H, Wang Y, Cheng Y, Yang H. The Study of Subthalamic Deep Brain Stimulation for Parkinson Disease-Associated Camptocormia. Med Sci Monit. 29 marzo 2020;26:e919682.

70. Nampiaparampil DE, Kuppy JE, Nampiaparampil GM, Salles SS. Inpatient rehabilitation after deep brain stimulator placement: a case series. Parkinsonism Relat Disord. 2008;14(4):356–8.

71. Canesi M, Lippi L, Rivaroli S, Vavassori D, Trenti M, Sartorio F, et al. Long-Term Impact of Deep Brain Stimulation in Parkinson’s Disease: Does It Affect Rehabilitation Outcomes? Medicina. 1 giugno 2024;60(6):927.

72. Armstrong JS, Forecasting LR. From crystal ball to computer. New York ua. 1985;348.

73. Roberts-Warrior D, Overby A, Jankovic J, Olson S, Lai EC, Krauss JK, et al. Postural control in Parkinson’s disease after unilateral posteroventral pallidotomy. Brain. 1 ottobre 2000;123(10):2141–9.

74. Rocchi L, Chiari L, Horak FB. Effects of deep brain stimulation and levodopa on postural sway in Parkinson’s disease. Journal of neurology, neurosurgery, and psychiatry. 2002;73(3):267–74.

75. Rocchi L, Chiari L, Cappello A, Gross A, Horak FB. Comparison between subthalamic nucleus and globus pallidus internus stimulation for postural performance in Parkinson’s disease. Gait & posture. aprile 2004;19(2):172–83.

76. Fasano A, Romito LM, Daniele A, Piano C, Zinno M, Bentivoglio AR, et al. Motor and cognitive outcome in patients with Parkinson’s disease 8 years after subthalamic implants. Brain. settembre 2010;133(9):2664–76.

77. St George RJ, Carlson-Kuhta P, Burchiel KJ, Hogarth P, Frank N, Horak FB. The effects of subthalamic and pallidal deep brain stimulation on postural responses in patients with Parkinson disease. Journal of neurosurgery. giugno 2012;116(6):1347–56.

78. St. George RJ, Nutt JG, Burchiel KJ, Horak FB. A meta-regression of the long-term effects of deep brain stimulation on balance and gait in PD. Neurology. 5 ottobre 2010;75(14):1292–9.

79. Hausdorff JM, Schaafsma JD, Balash Y, Bartels AL, Gurevich T, Giladi N. Impaired regulation of stride variability in Parkinson’s disease subjects with freezing of gait. Experimental brain research. 2003;149(2):187–94.

80. Xie T, Vigil J, MacCracken E, Gasparaitis A, Young J, Kang W, et al. Low-frequency stimulation of STN-DBS reduces aspiration and freezing of gait in patients with PD. Neurology. 2015;84(4):415–20.

81. Moreau C, Defebvre L, Destée A, Bleuse S, Clement F, Blatt JL, et al. STN-DBS frequency effects on freezing of gait in advanced Parkinson disease. Neurology. 8 luglio 2008;71(2):80–4.

82. van Nuenen BFL, Esselink RAJ, Munneke M, Speelman JD, van Laar T, Bloem BR. Postoperative gait deterioration after bilateral subthalamic nucleus stimulation in Parkinson’s disease. Mov Disord. 15 dicembre 2008;23(16):2404–6.

83. Weaver FM, Follett K, Stern M, Hur K, Harris C, Marks WJ, et al. Bilateral deep brain stimulation vs best medical therapy for patients with advanced Parkinson disease: a randomized controlled trial. JAMA. 7 gennaio 2009;301(1):63–73.

84. Bloem BR, Hausdorff JM, Visser JE, Giladi N. Falls and freezing of gait in Parkinson’s disease: a review of two interconnected, episodic phenomena. Movement disorders=:J: official journal of the Movement Disorder Society. agosto 2004;19(8):871–84.

85. Buhmann C, Huckhagel T, Engel K, Gulberti A, Hidding U, Poetter-Nerger M, et al. Adverse events in deep brain stimulation: A retrospective long-term analysis of neurological, psychiatric and other occurrences. PloS one [Internet]. 1 luglio 2017 [citato 1 maggio 2022];12(7). Disponibile su: https://pubmed.ncbi.nlm.nih.gov/28678830/

86. Dibble LE, Foreman KB, Addison O, Marcus RL, LaStayo PC. Exercise and Medication Effects on Persons With Parkinson Disease Across the Domains of Disability: A Randomized Clinical Trial. Journal of Neurologic Physical Therapy. aprile 2015;39(2):85.

87. Guidetti M, Bertini A, Pirone F, Sala G, Signorelli P, Ferrarese C, et al. Neuroprotection and Non-Invasive Brain Stimulation: Facts or Fiction? International Journal of Molecular Sciences 2022, Vol 23, Page 13775. 9 novembre 2022;23(22):13775.

88. Wallace BA, Ashkan K, Heise CE, Foote KD, Torres N, Mitrofanis J, et al. Survival of midbrain dopaminergic cells after lesion or deep brain stimulation of the subthalamic nucleus in MPTP-treated monkeys. Brain=:J: a journal of neurology. 2007;130(Pt 8):2129–45.

89. Shaw VE, Keay KA, Ashkan K, Benabid AL, Mitrofanis J. Dopaminergic cells in the periaqueductal grey matter of MPTP-treated monkeys and mice; patterns of survival and effect of deep brain stimulation and lesion of the subthalamic nucleus. Parkinsonism & related disorders. giugno 2010;16(5):338–44.

90. Harnack D, Meissner W, Jira JA, Winter C, Morgenstern R, Kupsch A. Placebo-controlled chronic high-frequency stimulation of the subthalamic nucleus preserves dopaminergic nigral neurons in a rat model of progressive Parkinsonism. Experimental neurology. marzo 2008;210(1):257–60.

91. Spieles-Engemann AL, Behbehani MM, Collier TJ, Wohlgenant SL, Steece-Collier K, Paumier K, et al. Stimulation of the rat subthalamic nucleus is neuroprotective following significant nigral dopamine neuron loss. Neurobiology of disease. luglio 2010;39(1):105–15.

92. Spieles-Engemann AL, Steece-Collier K, Behbehani MM, Collier TJ, Wohlgenant SL, Kemp CJ, et al. Subthalamic Nucleus Stimulation Increases Brain Derived Neurotrophic Factor in the Nigrostriatal System and Primary Motor Cortex. Journal of Parkinson’s disease. 5 maggio 2011;1(1):123.

93. Duncan RP, Van Dillen LR, Garbutt JM, Earhart GM, Perlmutter JS. Physical therapy and deep brain stimulation in Parkinson’s Disease: Protocol for a pilot randomized controlled trial. Pilot and Feasibility Studies. 25 aprile 2018;4(1):1–7.

94. Bötzel K, Kraft E. Strategies for treatment of gait and posture associated deficits in movement disorders: the impact of deep brain stimulation. Restor Neurol Neurosci. 2010;28(1):115–22.

95. Allert N, Cheeran B, Deuschl G, Barbe MT, Csoti I, Ebke M, et al. Postoperative rehabilitation after deep brain stimulation surgery for movement disorders. Clinical Neurophysiology. 1 marzo 2018;129(3):592–601.

96. Tomlinson CL, Herd CP, Clarke CE, Meek C, Patel S, Stowe R, et al. Physiotherapy for Parkinson’s disease: a comparison of techniques. Cochrane Database Syst Rev. 17 giugno 2014;2014(6):CD002815.

97. Monaco S, Renzi A, Galluzzi B, Mariani R, Di Trani M. The Relationship between Physiotherapist and Patient: A Qualitative Study on Physiotherapists’ Representations on This Theme. Healthcare (Basel). 25 ottobre 2022;10(11):2123.

98. Munhoz RP, Picillo M, Fox SH, Bruno V, Panisset M, Honey CR, et al. Eligibility Criteria for Deep Brain Stimulation in Parkinson’s Disease, Tremor, and Dystonia. Can J Neurol Sci. luglio 2016;43(4):462–71.

99. Yitayeh A, Teshome A. The effectiveness of physiotherapy treatment on balance dysfunction and postural instability in persons with Parkinson’s disease: A systematic review and meta-analysis. BMC Sports Science, Medicine and Rehabilitation. 11 febbraio 2016;8(1):1–10.

100. Radder DLM, Lígia Silva de Lima A, Domingos J, Keus SHJ, van Nimwegen M, Bloem BR, et al. Physiotherapy in Parkinson’s Disease: A Meta-Analysis of Present Treatment Modalities. Neurorehabil Neural Repair. ottobre 2020;34(10):871–80.

101. Angelopoulou E, Anagnostouli M, Chrousos GP, Bougea A. Massage therapy as a complementary treatment for Parkinson’s disease: A Systematic Literature Review. Complement Ther Med. marzo 2020;49:102340.

102. Bissolotti L, Isacco-Grassi F, Orizio C, Gobbo M, Berjano P, Villafañe JH, et al. Spinopelvic balance and body image perception in Parkinson’s disease: analysis of correlation. European Spine Journal. 6 ottobre 2015;24(7):898–905.

103. Ferrazzoli D, Ortelli P, Madeo G, Giladi N, Petzinger GM, Frazzitta G. Basal ganglia and beyond: The interplay between motor and cognitive aspects in Parkinson’s disease rehabilitation. Neuroscience & Biobehavioral Reviews. 1 luglio 2018;90:294–308.

104. Lageman SK, Mickens MN, Cash TV. Caregiver-identified needs and barriers to care in Parkinson’s disease. Geriatric Nursing. 1 maggio 2015;36(3):197–201.

105. Carter JH, Stewart BJ, Archbold PG, Inoue I, Jaglin J, Lannon M, et al. Living with a person who has parkinson’s disease: The Spouse’s perspective by stage of disease. Movement Disorders. 1998;13(1):20–8.

106. Dietz V. Proprioception and locomotor disorders. Nat Rev Neurosci. ottobre 2002;3(10):781– 90.

107. Kitago T, Krakauer JW. Chapter 8 - Motor learning principles for neurorehabilitation. In: Barnes MP, Good DC, curatori. Handbook of Clinical Neurology [Internet]. Elsevier; 2013 [citato 15 luglio 2024]. p. 93–103. (Neurological Rehabilitation; vol. 110). Disponibile su: https://www.sciencedirect.com/science/article/pii/B9780444529015000083

108. Defer GL, Widner H, Marié RM, Rémy P, Levivier M. Core assessment program for surgical interventional therapies in Parkinson’s disease (CAPSIT-PD). Mov Disord. luglio 1999;14(4):572–84.

109. Pollak P. Deep brain stimulation for Parkinson’s disease - patient selection. Handb Clin Neurol. 2013;116:97–105.

110. Jakobs M, Fomenko A, Lozano AM, Kiening KL. Cellular, molecular, and clinical mechanisms of action of deep brain stimulation-a systematic review on established indications and outlook on future developments. EMBO Mol Med. aprile 2019;11(4):e9575.

111. Jitkritsadakul O, Bhidayasiri R, Kalia SK, Hodaie M, Lozano AM, Fasano A. Systematic review of hardware-related complications of Deep Brain Stimulation: Do new indications pose an increased risk? Brain Stimulation. 1 settembre 2017;10(5):967–76.

112. Hamani C, Lozano AM. Hardware-related complications of deep brain stimulation: a review of the published literature. Stereotact Funct Neurosurg. 2006;84(5–6):248–51.

113. Deuschl G, Schade-Brittinger C, Krack P, Volkmann J, Schäfer H, Bötzel K, et al. A randomized trial of deep-brain stimulation for Parkinson’s disease. N Engl J Med. 31 agosto 2006;355(9):896–908.

114. Priori A, Maiorana N, Dini M, Guidetti M, Marceglia S, Ferrucci R. Adaptive deep brain stimulation (aDBS). International review of neurobiology. 2021;159:111–27.

115. Atkins D, Eccles M, Flottorp S, Guyatt GH, Henry D, Hill S, et al. Systems for grading the quality of evidence and the strength of recommendations I: Critical appraisal of existing approaches The GRADE Working Group. BMC Health Serv Res. 22 dicembre 2004;4(1):38.

116. Chudyk AM, Jutai JW, Petrella RJ, Speechley M. Systematic Review of Hip Fracture Rehabilitation Practices in the Elderly. Archives of Physical Medicine and Rehabilitation. 1 febbraio 2009;90(2):246–62.

117. O’Connor SR, Tully MA, Ryan B, Bradley JM, Baxter GD, McDonough SM. Failure of a numerical quality assessment scale to identify potential risk of bias in a systematic review: a comparison study. BMC Res Notes. 6 giugno 2015;8:224.

