## Supplementary Materials for "Physical therapy in patients with Parkinson’s disease treated with Deep Brain Stimulation: a Delphi panel study"

**Table 1. Modified Downs and Black Quality Assessment Checklist** [1]

| Article Title and Date: | | |
| --- | --- | --- |
| Authors: | | |
| # | Description | Answer (circle) |
| **REPORTING** | | |
| 1 | Is the hypothesis/aim/objective of the study clearly described? | Yes = 1  No = 0 |
| 2 | Are the main outcomes to be measured clearly described in the Introduction or Methods section? | Yes = 1  No = 0 |
| 3 | Are the characteristics of the patients included in the study clearly described? | Yes = 1  No = 0 |
| 4 | Are the main findings of the study clearly described? | Yes = 1  No = 0 |
| 5 | Does the study provide estimates of the random variability in the data for the main outcomes? | Yes = 1  No = 0 |
| 6 | Have the characteristics of patients lost to follow-up been described? | Yes = 1  No = 0 |
| 7 | Have actual probability values been reported (e.g. 0.035 rather than  <0.05) for the main outcomes except where the probability value is less than 0.001? | Yes = 1  No = 0 |
| **EXTERNAL VALIDITY** | | |
| 8 | Were the subjects asked to participate in the study representative of the entire population from which they were recruited? | Yes = 1  No/Unclear = 0 |
| 9 | Were those subjects who were prepared to participate representative of the entire population from which they were recruited? | Yes = 1  No/Unclear = 0 |
| **INTERNAL VALIDITY – BIAS** | | |
| 10 | If any of the results of the study were based on “data dredging”, was  this made clear? | Yes = 1  No/Unclear = 0 |
| 11 | Were the statistical tests used to assess the main outcomes appropriate? | Yes = 1  No/Unclear = 0 |
| 12 | Were the main outcome measures used accurate (valid and reliable)? | Yes = 1  No/Unclear = 0 |
| **INTERNAL VALIDITY – CONFOUNDING (SELECTION BIAS)** | | |
| 13 | Were losses of patients to follow-up considered? | Yes = 1  No/Unclear = 0 |
| Assessing the quality: excellent (11-13), good (9-10), fair (7-8), poor (≤6) [2,3] | | |

**Table 2. Questionnaire development as result of the systematic scoping review and the European Physiotherapy Guideline for Parkinson’s Disease** [4]

| **Benefit of PT** | **# Statement*** | **Reference** |
| --- | --- | --- |
| Motor symptoms and motor decline | S1, S5, S6 | [5–13] |
| Gait performance | S3 | [6–9,11,14,15] |
| Balance and postural instability | S2 | [5–8,11,12,14] |
| Quality of life or activities of daily living | S4 | [5,10–13,16] |
| Timing of PT treatment | S8, S9 | [5–9,15] |
| **PT treatment** | **# Statement** | **Reference** |
| Conventional physiotherapy | S12, S13 | [4,5,7,8,11,13–15] |
| Treadmill training | S14, S15 | [4,6,8,9,15,15] |
| Massage or Manual Therapy | S16, S17 | [4] |
| Cueing (visual, auditory) | S18, S19 | [4–6] |
| Dance-based training | S20, S21 | [4] |
| Tai Chi-based training | S22, S23 | [4] |
| Cognitive movement strategies | S24, S25 | [4] |
| Aerobic training | S26, S27 | [4,5,7,8] |
| Muscle strengthening | S28, S29 | [4,5,7,8,11,15] |
| Robot-assisted gait training | S30, S31 | [4] |
| Aquatic exercise | S32, S33 | [4] |
| Virtual reality and exergames | S34, S35 | [4] |
| Resistance training | S36, S37 | [4] |
| Exercise to improve trunk and limbs flexibility and range of motion | S38, S39 | [4,5,8,11] |

*S7, S10 AND S11 were added on the recommendation of Steering Committee as further aspects to be investigated

**REFERENCE**

[1]	Price OJ, Sewry N, Schwellnus M, Backer V, Reier-Nilsen T, Bougault V, et al. Prevalence of lower airway dysfunction in athletes: a systematic review and meta-analysis by a subgroup of the IOC consensus group on ‘acute respiratory illness in the athlete.’ Br J Sports Med 2022;56:213–22. https://doi.org/10.1136/bjsports-2021-104601.

[2]	Chudyk AM, Jutai JW, Petrella RJ, Speechley M. Systematic Review of Hip Fracture Rehabilitation Practices in the Elderly. Archives of Physical Medicine and Rehabilitation 2009;90:246–62. https://doi.org/10.1016/j.apmr.2008.06.036.

[3]	O’Connor SR, Tully MA, Ryan B, Bradley JM, Baxter GD, McDonough SM. Failure of a numerical quality assessment scale to identify potential risk of bias in a systematic review: a comparison study. BMC Res Notes 2015;8:224. https://doi.org/10.1186/s13104-015-1181-1.

[4]	Keus S, Munneke M, Graziano M, Paltamaa J, Pelosin E, Domingos J, et al. European Physiotherapy Guideline for Parkinson’s Disease Developed with twenty European professional associations 2014.

[5]	Tassorelli C, Buscone S, Sandrini G, Pacchetti C, Furnari A, Zangaglia R, et al. The role of rehabilitation in deep brain stimulation of the subthalamic nucleus for Parkinson’s disease: a pilot study. Parkinsonism Relat Disord 2009;15:675–81. https://doi.org/10.1016/j.parkreldis.2009.03.006.

[6]	Naro A, Pignolo L, Sorbera C, Latella D, Billeri L, Manuli A, et al. A Case-Controlled Pilot Study on Rhythmic Auditory Stimulation-Assisted Gait Training and Conventional Physiotherapy in Patients With Parkinson’s Disease Submitted to Deep Brain Stimulation. Front Neurol 2020;11:794. https://doi.org/10.3389/fneur.2020.00794.

[7]	Bestaven E, Guillaud E, De Sèze M, Jerome A, Burbaud P, Cazalets J-R, et al. Effect of Trunk Muscle Strengthening on Gait Pattern and Falls in Parkinson’s Disease. J Rehabil Med Clin Commun 2019;2:1000003. https://doi.org/10.2340/20030711-1000003.

[8]	Canesi M, Lippi L, Rivaroli S, Vavassori D, Trenti M, Sartorio F, et al. Long-Term Impact of Deep Brain Stimulation in Parkinson’s Disease: Does It Affect Rehabilitation Outcomes? Medicina 2024;60:927. https://doi.org/10.3390/medicina60060927.

[9]	Nardo A, Anasetti F, Servello D, Porta M. Quantitative gait analysis in patients with Parkinson treated with deep brain stimulation: the effects of a robotic gait training. NeuroRehabilitation 2014;35:779–88. https://doi.org/10.3233/NRE-141173.

[10]	Cohen DB, Oh MY, Baser SM, Angle C, Whiting A, Birk C, et al. Fast-track programming and rehabilitation model: a novel approach to postoperative deep brain stimulation patient care. Arch Phys Med Rehabil 2007;88:1320–4. https://doi.org/10.1016/j.apmr.2007.06.770.

[11]	Sato K, Aita N, Hokari Y, Kitahara E, Tani M, Izawa N, et al. Balance and Gait Improvements of Postoperative Rehabilitation in Patients with Parkinson’s Disease Treated with Subthalamic Nucleus Deep Brain Stimulation (STN-DBS). Parkinsons Dis 2019;2019:7104071. https://doi.org/10.1155/2019/7104071.

[12]	Li H, Liang S, Yu Y, Wang Y, Cheng Y, Yang H, et al. Clinical experience of comprehensive treatment on the balance function of Parkinson’s disease. Medicine (Baltimore) 2020;99:e20154. https://doi.org/10.1097/MD.0000000000020154.

[13]	Liang S, Yu Y, Li H, Wang Y, Cheng Y, Yang H. The Study of Subthalamic Deep Brain Stimulation for Parkinson Disease-Associated Camptocormia. Med Sci Monit 2020;26:e919682. https://doi.org/10.12659/MSM.919682.

[14]	Sato K, Hokari Y, Kitahara E, Izawa N, Hatori K, Honaga K, et al. Short-Term Motor Outcomes in Parkinson’s Disease after Subthalamic Nucleus Deep Brain Stimulation Combined with Post-Operative Rehabilitation: A Pre-Post Comparison Study. Parkinsons Dis 2022;2022:8448638. https://doi.org/10.1155/2022/8448638.

[15]	Luna NMS, Lucareli PRG, Sales VC, Speciali D, Alonso AC, Peterson MD, et al. Treadmill training in Parkinson’s patients after deep brain stimulation: Effects on gait kinematic. NeuroRehabilitation 2018;42:149–58. https://doi.org/10.3233/NRE-172267.

[16]	Nampiaparampil DE, Kuppy JE, Nampiaparampil GM, Salles SS. Inpatient rehabilitation after deep brain stimulator placement: a case series. Parkinsonism Relat Disord 2008;14:356–8. https://doi.org/10.1016/j.parkreldis.2007.07.002.
